# Transcriptional profiles predict treatment outcome in patients with tuberculosis and diabetes at diagnosis and at two weeks after initiation of anti-tuberculosis treatment

**DOI:** 10.1101/2022.02.08.22269796

**Authors:** Cassandra L.R. van Doorn, Clare Eckold, Katharina Ronacher, Rovina Ruslami, Suzanne van Veen, Ji-Sook Lee, Vinod Kumar, Sarah Kerry-Barnard, Stephanus T. Malherbe, Léanie Kleynhans, Kim Stanley, Philip C. Hill, Simone A. Joosten, Reinout van Crevel, Cisca Wijmenga, Julia A. Critchley, Gerhard Walzl, Bachti Alisjahbana, Mariëlle C. Haks, Hazel M. Dockrell, Tom H. M. Ottenhoff, Eleonora Vianello, Jacqueline M. Cliff, the TANDEM Consortium

## Abstract

**Background:** Globally, the anti-tuberculosis (TB) treatment success rate is approximately 85%, with treatment failure, relapse and death occurring in a significant proportion of pulmonary TB patients. Treatment success is lower among people with diabetes mellitus (DM). Predicting treatment failure early after diagnosis would allow early treatment adaptation and may improve global TB control.

**Methods:** Samples were collected in a longitudinal cohort study of adult TB patients with or without concomitant DM from South Africa and Indonesia to characterize whole blood transcriptional profiles before and during anti-TB treatment, using unbiased RNA-Seq and targeted gene dcRT-MLPA.

**Findings:** We report differences in whole blood transcriptome profiles, which were observed before initiation of treatment and throughout treatment, between patients with a good versus poor anti-TB treatment outcome. An eight-gene and a 22-gene blood transcriptional signature distinguished patients with a good treatment outcome from patients with a poor treatment outcome at diagnosis (AUC=0·815) or two weeks (AUC=0·834) after initiation of anti-TB treatment, respectively. High accuracy was obtained by cross-validating this signature in an external cohort (AUC=0·749).

**Interpretation:** These findings suggest that transcriptional profiles can be used as a prognostic biomarker for treatment failure and success, even in patients with concomitant DM.

**Funding:** The research leading to these results, as part of the TANDEM Consortium, received funding from the European Community’s Seventh Framework Programme (FP7/2007-2013 Grant Agreement No. 305279) and the Netherlands Organization for Scientific Research (NWO-TOP Grant Agreement No. 91214038).

## Introduction

With more than 10 million new cases and approximately 1·5 million deaths annually, tuberculosis (TB), which is caused by *Mycobacterium tuberculosis* (*Mtb*), continues to be a major global health threat.^1^ Upon infection with *Mtb*, 5-10% of adults develop active disease during their lifetime and one quarter of the world’s population is estimated to be latently infected with *Mtb* (LTBI). The global anti-TB treatment success rate is only about 85% and even lower in patients with multi-drug resistant TB or with comorbidities like HIV or diabetes mellitus (DM)^1–3^, resulting in a significant number of patients with poor clinical outcomes.

DM triples the risk of developing active TB^4^ and increases the risk of poor clearance of the infection following anti-TB treatment.^5–7^ In 2020, 0·37 million TB cases were estimated to suffer from DM comorbidity.^1^ 85-95% of all DM cases is attributed to type-2 diabetes mellitus (T2DM).^8^ Since global DM prevalence is estimated to rise from 463 million people in 2019 to 700 million in 2045^9^, in particular in areas where TB is endemic, there is increasing concern about the consequences of the rising DM prevalence for global TB control.^1^ The mechanisms underlying DM-induced treatment failure remain, however, poorly understood.

Prediction of treatment failure based on sputum-smear microscopy and mycobacterial culture lacks sensitivity^10^ and depends on the quality of sputum samples, which are difficult to collect and are frequently inconsistent in quality.^11–13^ In addition to more advanced sputum-based diagnostics, monitoring of whole blood transcriptomics may be an additional, complementary but independent method to monitor treatment responses, possibly with increased sensitivity.^14^ Numerous studies have reported transcriptional biomarker profiles for active TB and response to anti-TB treatment using whole-blood or PBMCs in settings with varying TB incidence.^15–20^ In addition, multiple studies have demonstrated the predictive potential of host gene biomarkers in identifying patients at risk of developing active TB, relapse and treatment failure.^21–28^ Together, these studies showed that gene signatures may have utility at predicting anti-TB treatment success versus failure already early after TB diagnosis, providing a significant improvement over the currently used low sensitivity conversion to negative sputum-based culture testing.^10^ Despite the high incidence of DM and pre-DM among TB patients in TB-endemic settings^7, 29–31^, only a few studies have identified or validated such signatures in TB patients with DM or hyperglycemia.^32, 33^

Characterizing transcriptomic profiles may improve our understanding towards the immunological pathways that are involved in DM-associated TB pathology and monitoring treatment success and failure in TB patients with concomitant DM is key to combat the tuberculosis-diabetes (TB-DM) co-epidemic. Although the blood transcriptome profile of TB-DM patients is more similar to TB patients than to DM patients, suggesting a dominant influence of active TB infection, we and others recently demonstrated significant differences in the blood transcriptome of TB-DM patients compared to TB patients.^32, 33^ Additionally, the transcriptomic profiles of patients with TB-related intermediate hyperglycemia (TBrel-IH) were similar to the profiles of TB-DM patients.^32^ Importantly, we also showed that DM comorbidity lowered the performance of published diagnostic biomarker signatures.^32^ Therefore, there is a need for biomarkers that predict treatment success and failure in TB patients independently of their glycemia or DM status.

The aim of the current study was to identify blood transcriptional gene signature for predicting the anti-TB treatment outcome at an early stage after initiation of anti-TB treatment, irrespective of concomitant DM. We combined an unbiased RNA-Seq approach and a selective dcRT-MLPA approach (a multiplex RT-PCR platform) as two independent strategies to identify gene signatures with high discriminatory power to distinguish patients with a good treatment outcome from patients with a poor treatment outcome. Host gene biomarker profiles to identify anti-TB treatment success or failure could facilitate the evaluation of new anti-TB drugs and improve clinical surveillance of TB patients, even in settings with high DM incidence.

## Methods

### Study participant recruitment, classification and treatment

Adult pulmonary TB patients were recruited as part of the TANDEM project^29^ in two locations: Bandung in Indonesia (UNPAD) and Cape Town in South Africa (SUN). All TB patients were newly diagnosed and microbiologically confirmed, and included people with TB-DM. The TB-DM group included participants with both pre-diagnosed DM and newly identified DM, with new diagnosis based on a laboratory HbA1c test ≥6·5% with a confirmatory HbA1c test ≥6·5% or fasting blood glucose ≥7 mmol/L at TB diagnosis^29^, followed by a further HbA1c test ≥6·5% after 6 months of TB treatment. The TB patients without DM included people with a normal glycaemic index (laboratory HbA1c <5·7%) at TB diagnosis (“TB-only”). Patients whose HbA1c test results were ≥5.7% and <6.5% at both TB diagnosis and at 6 months were deemed to have pre-diabetes (“TB-preDM”) and patients with raised glycaemia at TB diagnosis but below the cut-off for DM diagnosis (5·7%≤ laboratory HbA1c <6·5%) were deemed to have TB-related intermediate hyperglycaemia (“TBrel-IH”). In South Africa, healthy controls without TB or DM were also recruited for baseline sample analysis. Multi-drug-resistant TB, HIV positivity, pregnancy, serious co-morbidity and corticosteroid use were exclusion criteria. TB patients received standard first line TB treatment according to WHO Guidelines.

Microbiological measures recorded at baseline and throughout treatment included sputum smear and culture, with time to positivity (TTP) in mycobacteria growth indicator tubes (MGIT) also assayed in South Africa. TB patients were classified based on their treatment outcome: “poor treatment outcome” included those patients who died, failed initial treatment (remained sputum positive at five months) or experienced TB-recurrence in the 12 month follow-up period post treatment, whilst those with “good treatment outcome” had successful TB treatment without subsequent recurrence. Patients for whom the outcome data were missing were not included in downstream analyses. Most TB-DM patients received local standard of care DM treatment, whilst a subgroup in Indonesia received more intensive HbA1c monitoring and treatment adjustment through TB treatment as part of a pragmatic randomised control trial.^34^

### Ethics Statement

The study was approved by the London School of Hygiene & Tropical Medicine Observational Research Ethics Committee (6449), the SUN Health Research Ethics Committee (N13/05/064) and the UNPAD Health Research Ethics Committee, Faculty of Medicine, Universitas Padjadjaran (number 377/UN6.C2.1.2/ KEPK/ PN), and participants gave written informed consent.

### RNA sample collection and extraction

Patient samples were collected prior to initiation of treatment (diagnosis), at weeks 2, 4, 8, 16 and 26 through treatment, and at 12 months after TB diagnosis, and from HC at baseline only. Venous blood (2·5ml) was collected into PAXgene Blood RNA Tubes (PreAnalytiX). Total RNA was extracted using RNeasy spin columns (Qiagen) and quantified by Nanodrop (Agilent). The LabChip GX HiSens RNA system (PerkinElmer) was used for quality assessment of samples processed by RNA-Seq.

### Unbiased RNA-Seq of global gene expression

Samples collected at TB diagnosis and weeks 2, months 2, and months 6 from the first 63 participants recruited were analysed by RNA-Seq (Table 1). Libraries were generated using the poly-A tail Bioscientific NEXTflex-Rapid-Directional mRNA-Seq method with the Caliper SciClone. Single-end sequencing was performed using the NextSeq500 High Output kit V2 (Illumina) for 75 cycles. Sequence data from FASTQ files were aligned to the Human g1kv37 reference genome, using STAR (v2.5.1b).^35^ Quality control was performed with FastQC^36^, while transcript quantification was performed using HT-seq count (v0.61)^37^: lowly expressed transcripts (<50 counts across all samples), were removed from the downstream analysis. RNA-Seq data were normalised using DESeq2 (v1.30.0).^38^

**Table 1.**
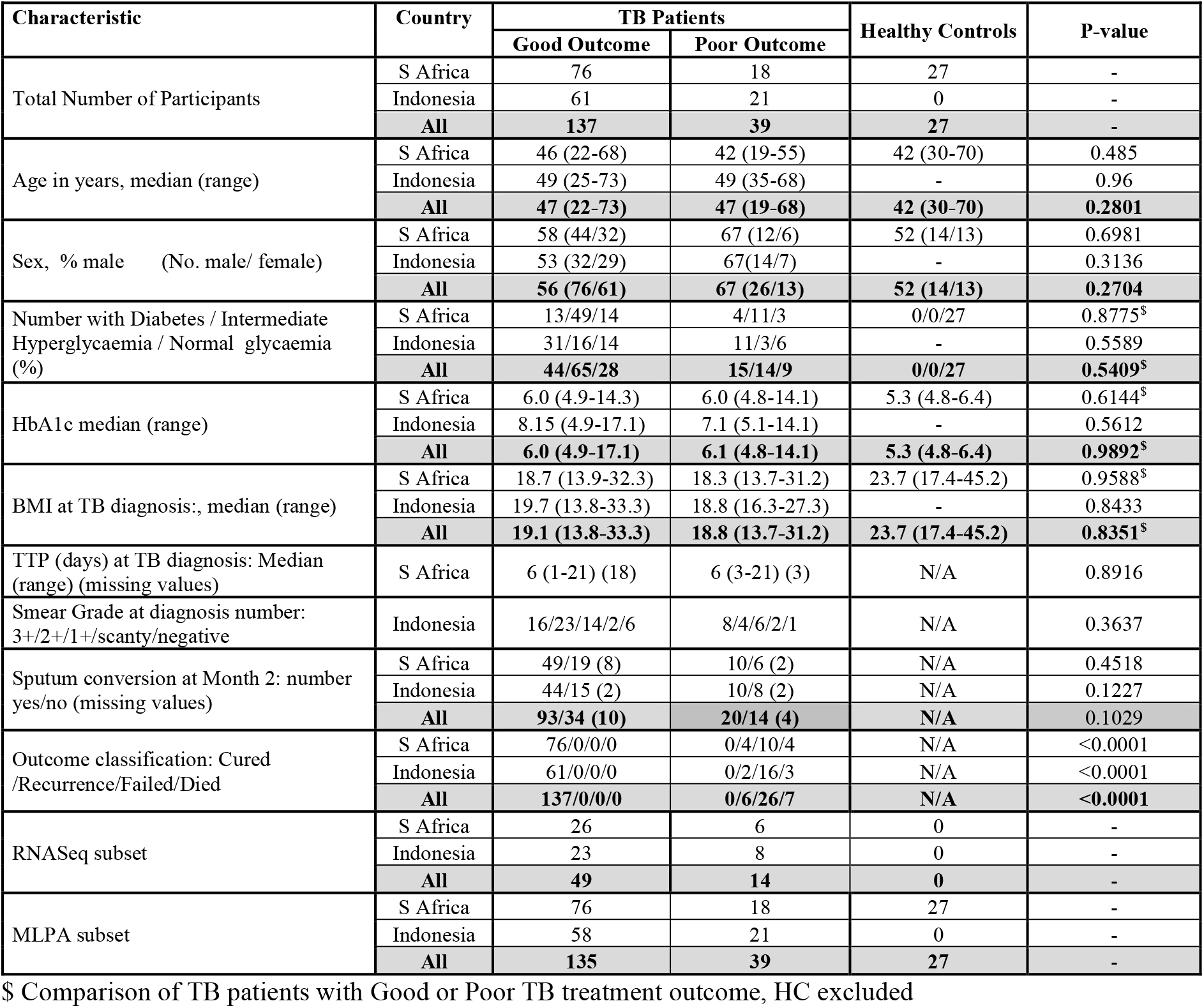
Study Participant Demographics

### Dual-color Reverse-Transcriptase Multiplex Ligation-dependent Probe Amplification (dcRT-MLPA)

Dual-color reverse-Transcriptase Multiplex Ligation-dependent Probe Amplification (dcRT-MLPA) was performed on all samples to identify blood transcriptional profiles as described previously.^39^ Brief descriptions are provided in the Supplementary Information. RT primers and half-probes were designed by Leiden University Medical Centre (LUMC, Leiden, The Netherlands) and encompassed sequences for 144 selected key immune-related genes to profile the innate, adaptive and inflammatory immune responses (Supplementary Table S1), and four housekeeping genes (*GAPDH, ABR, GUSB, B2M*). Genes with an adjusted p-value <0·05 (Benjamini-Hochberg^40^) and a log2-fold change (FC) <-0·6 and >0·6 were considered differentially expressed genes (DEGs). Genes that were below the detection limit in >90% of the samples per cohort were excluded from analysis.

### Data analysis and Statistics

Statistical analyses to compare participant demographics were carried out using GraphPad Prism 8 software (Graphpad Software, San Diego, CA, USA). For continuous measures, a Mann-Whitney U-test was used when comparing two groups and a Kruskal-Wallis test when comparing three groups. For non-continuous measures, the Chi-square test was used. P-values as <0·05 were considered significant.

Molecular Degree of Perturbation (MDP) analysis was performed by R using *mdp* R package^41^, and differences between the mean ranks of the groups were assessed by Mann-Whitney U test followed by Benjamini-Hochberg False discovery correction.^40^ Cell population estimates were calculated using the cellCODE^42^ R package, with the IRIS^43^ and DMAP^44^ data sets used as a reference. Modular analysis was performed using the R package tmod^45^ and its HGtest method, with DEGs used as the foreground and all genes used as the background signal.

Differential expression analysis (DEA) was performed in R using the MaSigPro package^46^ to characterise longitudinal differential gene expression of genes measured by RNA-Seq: this followed a two-step regression method, finding genes with significant temporal expression changes and also significant differences between clinical groups. A quadratic regression model was executed due to the number of timepoints analysed. The regression model treats time as a quantitative variable so differentially expressed are not only detected, but also changes in trends and magnitude.

Longitudinal DEA of genes measured by dcRT-MLPA was assessed by means of linear mixed models for repeated measures over time using lme4 package in R.^47^ A Benjamini- Hochberg False discovery correction was performed, with an adjusted p-value of <0·05 deemed significant. Non-parametric Mann-Whitney U-test followed by Benjamini-Hochberg correction was performed to identify DEGs between patients who had a good and poor treatment outcome. Correlations were evaluated using Pearson’s correlation coefficient.

Treatment outcome signatures based on dcRT-MLPA data were identified in TB patients of South Africa and Indonesia using Recursive Feature Elimination (RFE)^48^ and Random Forest (RF). Because the number of patients with a good treatment outcome was considerably larger than those with a poor treatment outcome (poor, n=38; good, n=134), a random down-sampling technique as well as a Synthetic Minority Oversampling Technique (SMOTE) were applied to balance the classes (i.e. “good treatment outcome” and “poor treatment outcome”) of the dataset.^49^ RF was performed as machine learning algorithm on the dataset including the selected genes and the performance of gene signatures was evaluated by Leave-One-Out Cross Validation (LOOCV).^50, 51^ We assessed the classifying performance of the model by evaluating Receiver Operating Characteristic (ROC) curve and Area Under the ROC Curve (AUC) with 95% Confidence Interval (CI). An extended description of the data-analysis methods is provided in the Supplementary Information.

### Role of Funders

Funders had no role in study design, data collection, data analyses, data interpretation, writing of the report and decision to submit the paper for publication.

## Results

### Study Design and Cohort

Pulmonary TB patients were recruited into the prospective longitudinal study in South Africa and Indonesia, and followed up through standard treatment and for the following 12 months. Altogether, 39 TB patients of the 176 recruited had a “poor treatment outcome”, with 7 patients dying, 26 failing treatment (based on continued sputum smear or culture positivity at month 6), and 6 experiencing recurrences in the subsequent 18 months. The “poor treatment outcome” rates were similar in the two sites (Table 1). The median age of the patients was equal in patients with either a good or poor treatment outcome (median = 47 years), with a higher proportion of males with a poor treatment outcome than a good treatment outcome (56% and 67% respectively). The proportion of TB patients with DM with a poor treatment outcome (15/39; 38%) was slightly higher than that with a good treatment outcome (44/137; 32%), whereas the proportion with TBrel-IH was higher in those with a good treatment outcome (65/137; 47%) than a poor treatment outcome (14/39; 36%). There was no evidence that those who had a poor treatment outcome had more severe TB at diagnosis, with similar sputum bacterial loads (as measured by TTP) in TB patients from South Africa and similar sputum smear grade in Indonesia across the good and poor treatment outcome groups.

### Poor treatment outcome was reflected by an attenuated treatment response compared to good treatment outcome

The holistic unbiased analysis of gene expression in TB patients with good or poor treatment outcomes by RNA-Seq approach was performed on a subset of study participants (Table 1). There were significant changes in global gene expression in patients with a good treatment outcome continuously through TB treatment, reflecting treatment response (Figure 1A). Gene expression perturbation was also evident in patients who had a poor treatment outcome, although the sample score was higher at diagnosis compared to patients who had a good treatment outcome. This represents differences at the transcriptomic level between patients with a good versus a poor treatment outcome, already before initiation of anti-TB treatment. This was followed by less change over time in response to TB treatment in the poor TB outcome group.

**Figure 1.**
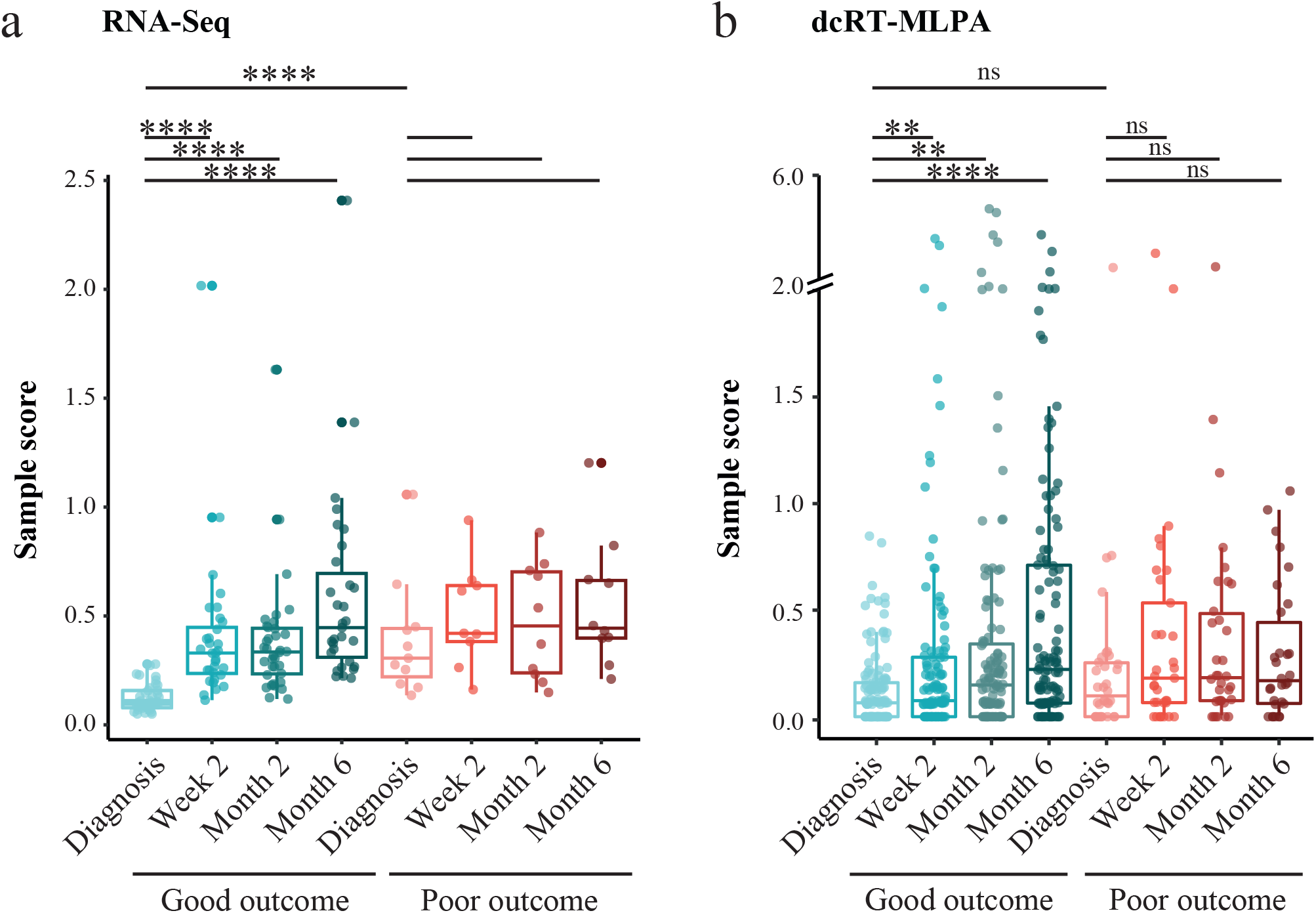
MDP plots representing the change in gene expression perturbation in TB patients categorized based on treatment outcome. Full blood transcriptomes from TB patients who had a good or poor treatment outcome were determined by (A) RNA-Seq and by (B) dcRT-MLPA. The extent of overall difference in gene expression, relative to the median of expression at diagnosis in those who had a good treatment outcome, was calculated for individual patients at the timepoints shown. The bars and whiskers show the median and data within the Q_1_-1·5 x inter quartile range (IQR) and Q_3_+1·5 x IQR interval. Differences were significant by Mann-Whitney U-test with Benjamini-Hochberg correction for multiple testing. * p<0·05, ** p<0·01, *** p<0·001, **** p<0·0001.

Next, we focused our molecular distance analysis on 144 TB-associated genes as measured by dcRT-MLPA, which was performed on all study participants (n=201) (Table 1). Again, there were significant changes in global gene expression continuously through TB treatment in patients with a good treatment outcome, but not in patients with a poor treatment outcome (Figure 1B), reflecting an attenuated TB treatment response. Despite the substantial treatment response in patients who had a good treatment outcome, gene expression perturbation did not completely normalize to levels of healthy controls by 6 months (Supplementary Figure S1).

Together, these data suggest that there was a different biosignature in those with good versus poor treatment outcomes, which was reflected by transcriptomic differences before initiation of anti-TB treatment and by a tempered response to anti-TB treatment in patients with a poor treatment outcome compared to patients with a good treatment outcome.

### Global Differential expression in patients who had good or poor responses to TB treatment

The changes in gene expression in the RNA-Seq data through time and between the patients with a good or poor treatment outcome were analyzed by MaSigPro, initially in the South African and Indonesian cohorts separately. The strength in this method was that it was able to monitor the change in dynamics over time and also between different treatment groups. In South Africa, the genes which changed differently through time between the patients with good or poor treatment outcome were grouped into nine clusters (Supplementary Figure S2A, Supplementary Table S2), with an increase in expression through treatment in four clusters (1,2,3,5), a decrease in four clusters (4,6,7,8), and no change in one cluster (9). Genes with higher expression throughout treatment in TB patients with poor treatment outcome were grouped in seven clusters (1,3,4,5,7,8,9) and those with higher expression in TB patients with a good treatment outcome were grouped in two clusters (2,6). A similar pattern was observed in the Indonesian cohort, with DEGs identified through treatment between good and poor separating into nine clusters (genes increasing in clusters 2,5,7, and decreasing in clusters 1,3,4,6,8,9) (Supplementary Figure S2B, Supplementary Table S3), with higher expression in either the good or poor treatment outcome group. Importantly, these differences in gene expression through time were observed in all TB patient groups, irrespective of their DM status. The genes grouped into the clusters in the Indonesian and South African cohorts partially overlapped but there was variability. A third MaSigPro analysis was therefore conducted, to find those genes which were differentially expressed through treatment between TB patients with a good or poor treatment outcome, irrespective of their geographical origin (Figure 2, Supplementary Table S4). Again, the genes differentially expressed through treatment in the combined analysis separated into nine clusters, with variable patterns of expression over time and between TB patients with good or poor treatment outcome. Some clusters (2,5,6) contained genes which were different between the groups at all time points, whereas other clusters (1,3,4,5,7,8,9) were similar at some timepoints and more divergent at others (Figure 2, Table 2).

**Figure 2.**
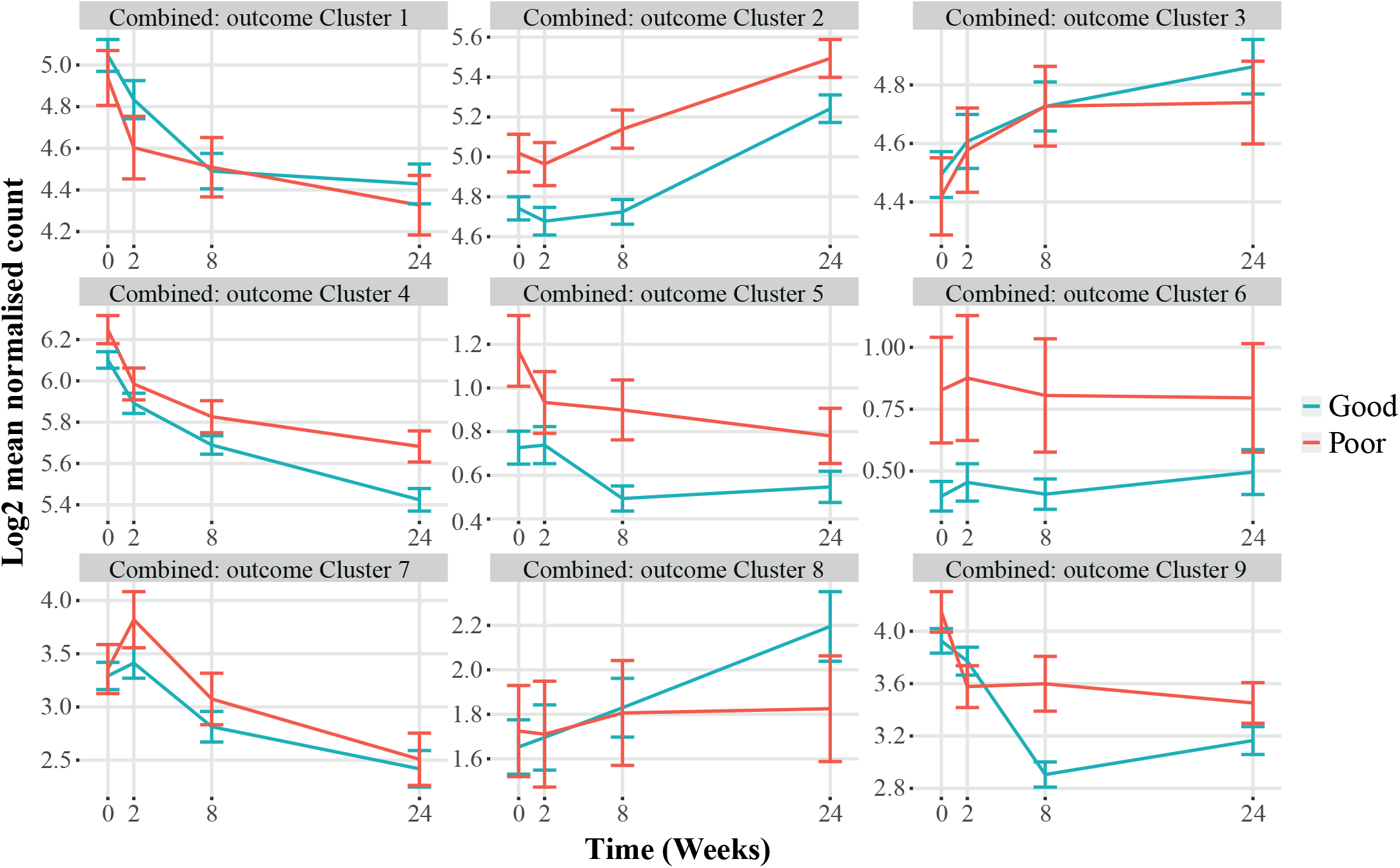
MaSigPro analysis of TB patients with good or poor treatment outcome, across combined South African and Indonesian cohorts. Plots show hierarchical clusters of genes in patients with a good (blue) or poor (red) treatment outcome. Bars show mean ± 1 SEM. Data were filtered to remove lowly abundant transcripts prior to analysis.

**Table 2.**
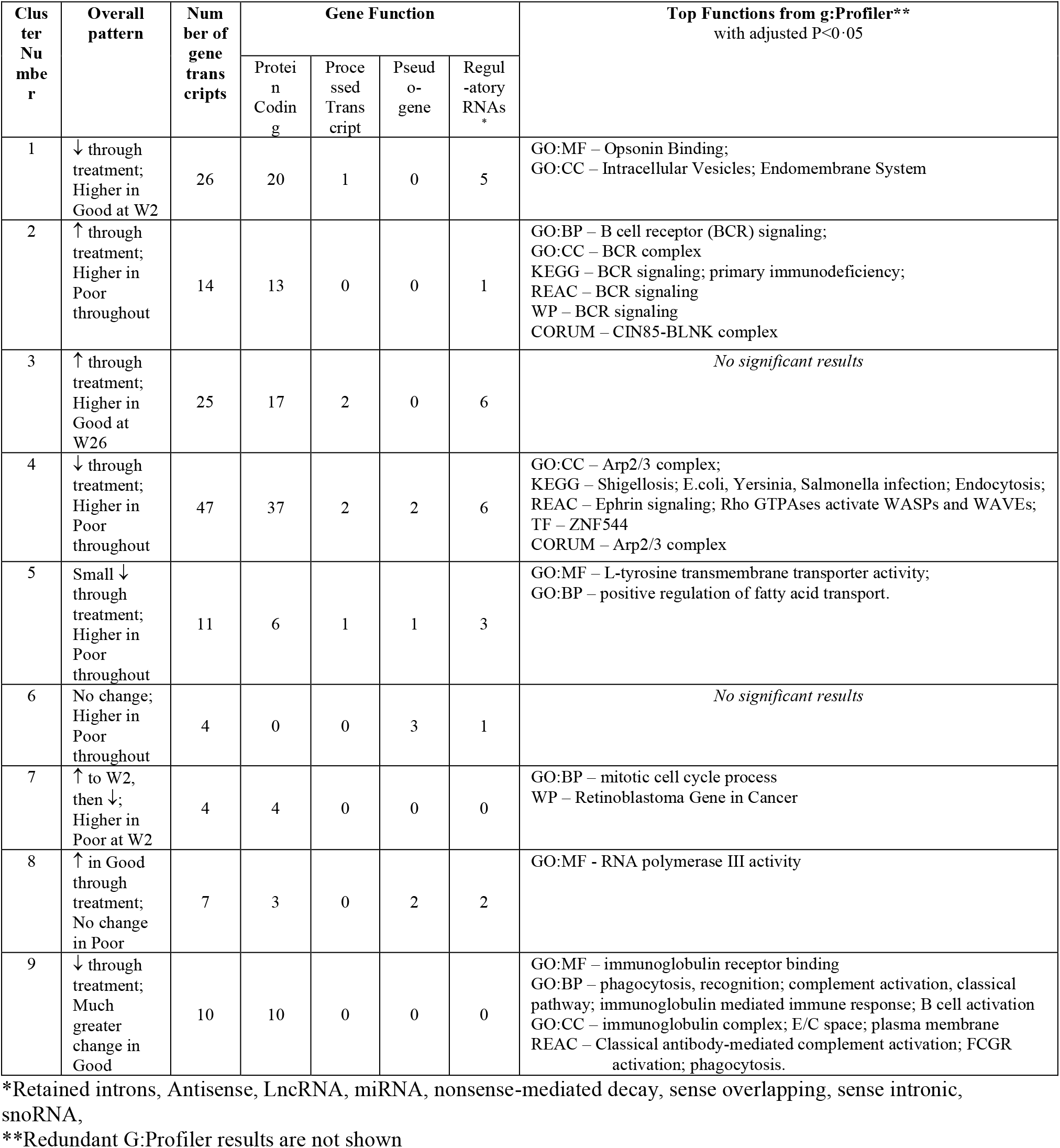
Clusters of genes differentially expressed between TB patients with good or poor treatment outcomes to TB treatment in MaSigPro analysis of combined RNA-Seq data from South Africa and Indonesia

| Cluster Number | Overall pattern | Number of gene transcripts | Gene Function |  |  |  | Top Functions from g:Profiler**<br>with adjusted P<0.05 |
| --- | --- | --- | --- | --- | --- | --- | --- |
|  |  |  | Protein Coding | Processed Transcript | Pseudogene | Regulatory RNAs* |  |
| 1 | ↓ through treatment;<br>Higher in Good at W2 | 26 | 20 | 1 | 0 | 5 | GO:MF – Opsonin Binding;<br>GO:CC – Intracellular Vesicles; Endomembrane System |
| 2 | ↑ through treatment;<br>Higher in Poor throughout | 14 | 13 | 0 | 0 | 1 | GO:BP – B cell receptor (BCR) signaling;<br>GO:CC – BCR complex<br>KEGG – BCR signaling; primary immunodeficiency;<br>REAC – BCR signaling<br>WP – BCR signaling<br>CORUM – CIN85-BLNK complex |
| 3 | ↑ through treatment;<br>Higher in Good at W26 | 25 | 17 | 2 | 0 | 6 | *No significant results* |
| 4 | ↓ through treatment;<br>Higher in Poor throughout | 47 | 37 | 2 | 2 | 6 | GO:CC – Arp2/3 complex;<br>KEGG – Shigellosis; E.coli, Yersinia, Salmonella infection; Endocytosis;<br>REAC – Ephrin signaling; Rho GTPases activate WASPs and WAVES;<br>TF – ZNF544<br>CORUM – Arp2/3 complex |
| 5 | Small ↓ through treatment;<br>Higher in Poor throughout | 11 | 6 | 1 | 1 | 3 | GO:MF – L-tyrosine transmembrane transporter activity;<br>GO:BP – positive regulation of fatty acid transport. |
| 6 | No change;<br>Higher in Poor throughout | 4 | 0 | 0 | 3 | 1 | *No significant results* |
| 7 | ↑ to W2,<br>then ↓;<br>Higher in Poor at W2 | 4 | 4 | 0 | 0 | 0 | GO:BP – mitotic cell cycle process<br>WP – Retinoblastoma Gene in Cancer |
| 8 | ↑ in Good through treatment;<br>No change in Poor | 7 | 3 | 0 | 2 | 2 | GO:MF – RNA polymerase III activity |
| 9 | ↓ through treatment;<br>Much greater change in Good | 10 | 10 | 0 | 0 | 0 | GO:MF – immunoglobulin receptor binding<br>GO:BP – phagocytosis, recognition; complement activation, classical pathway; immunoglobulin mediated immune response; B cell activation<br>GO:CC – immunoglobulin complex; E/C space; plasma membrane<br>REAC – Classical antibody-mediated complement activation; FCGR activation; phagocytosis. |

\*Retained introns, Antisense, LncRNA, miRNA, nonsense-mediated decay, sense overlapping, sense intronic, snoRNA,
\*\*Redundant G:Profiler results are not shown

The number of transcripts within each cluster in the combined analysis ranged from 4 to 47 (Table 2), with the majority of genes identified in all clusters encoding proteins. There were also various regulatory transcripts in some clusters, including long non-coding RNAs, miRNA, snoRNA, retained introns, as well as antisense, nonsense-mediated decay, overlapping senses and sense intronic transcripts. To understand the biological function of the DEGs, the transcripts within each cluster were analysed using the g:COST tool within the g:Profiler application^52^, to determine significant enrichment of genes in Gene Ontology (GO) molecular function, cellular component and biological process categories, as well as in curated biological pathways from KEGG and Reactome databases and the CORUM protein database. Genes in cluster 2 were largely involved in B cell receptor signalling, seen in the GO and pathway analyses, and these were more highly expressed in people who had a poor treatment outcome, with increasing expression through treatment. This upregulation of genes involved in B cell function, particularly those involved in earlier stages of B cell development, was not related to the overall number of B cells in the samples, as predicted from the samples using CellCode analysis package (Supplementary Figure S3). Cluster 9 was predominantly composed of immunoglobulin transcripts, whose expression decreased much more substantially in patients with a good treatment outcome. The largest gene cluster (4) was enriched with genes involved in actin remodelling, including the Arp 2/3 complex, and in pathways related to infections with bacteria such as Shigella, *E. coli*, Yersinia and Salmonella.

Cluster 7 contained genes related to mitotic cell division, and these were more highly expressed in patients with a poor treatment outcome (Table 2). These analyses were also performed using the DAVID online tool^53^, and similar results were obtained (not shown). The DEGs found in the combined and separate cohort MaSigPro analyses were used as a foreground against all genes in a modular analysis using the Tmod package, which gives biological function to a gene list. It showed an upregulation of genes involved in B cell function in good versus poor treatment outcomes, in both the Indonesian and South African cohorts (Supplementary Table S5).

### Identification of DEGs through TB treatment in patients with good or poor treatment outcomes

Next, we focused our DEA on 144 genes that previously have been associated with TB^39^ using dcRT-MLPA (Supplementary Table S6). No significant DEGs were detected by directly comparing patients with a good versus a poor treatment outcome at the indicated timepoints (Supplementary Figure S4), and therefore we analyzed longitudinal expression of genes. Kinetic profiling of DEGs identified 16 DEGs in patients with a good treatment outcome and 12 DEGs in patients with a poor treatment outcome. The longitudinal expression of DEGs identified by dcRT-MLPA showed a significant correlation with genes measured by RNA-Seq, highlighting the validity and reproducibility of our approach (Supplementary Figure S5). A high correlation between DEGs of patients who had a poor treatment outcome and DEGs of patients who had a good treatment outcome could be detected (R=0·87, p<0·0001), highlighting the challenge of discriminating patients with a good versus a poor treatment outcome based on single genes (Supplementary Figure S6). Genes associated with active TB^15, 20, 54^ or risk of developing TB^22^ were substantially downregulated (*GBP1, GBP2, GBP5,* and *IFITM3*) or upregulated (*GNLY* and *PRF1*) over time in TB patients regardless of their treatment outcome, reflecting transcriptomic response to anti-TB treatment (Figure 3A and Supplementary Figure S7). Other genes associated with active TB were significantly down- or upregulated (*STAT2, MMP9, IRF7, IFI6, IFIT2*, *IFIT3,* and *CCR7*) during anti-TB treatment in patients who had a good treatment outcome, but not in patients who had a poor treatment outcome, or vice versa (*CD3E, PTPRCv1, NLRP1, BCL2*)^15, 39, 54, 55^. The expression of *TAGAP,* previously associated with active TB^55^, was significantly increased during anti-TB treatment in patients who had a poor treatment outcome. Modular analysis showed that the gene profile of regulated genes was dominated by genes in the interferon (IFN) signaling pathway, especially in patients who had a good treatment outcome (Figure 3B).

**Figure 3.**
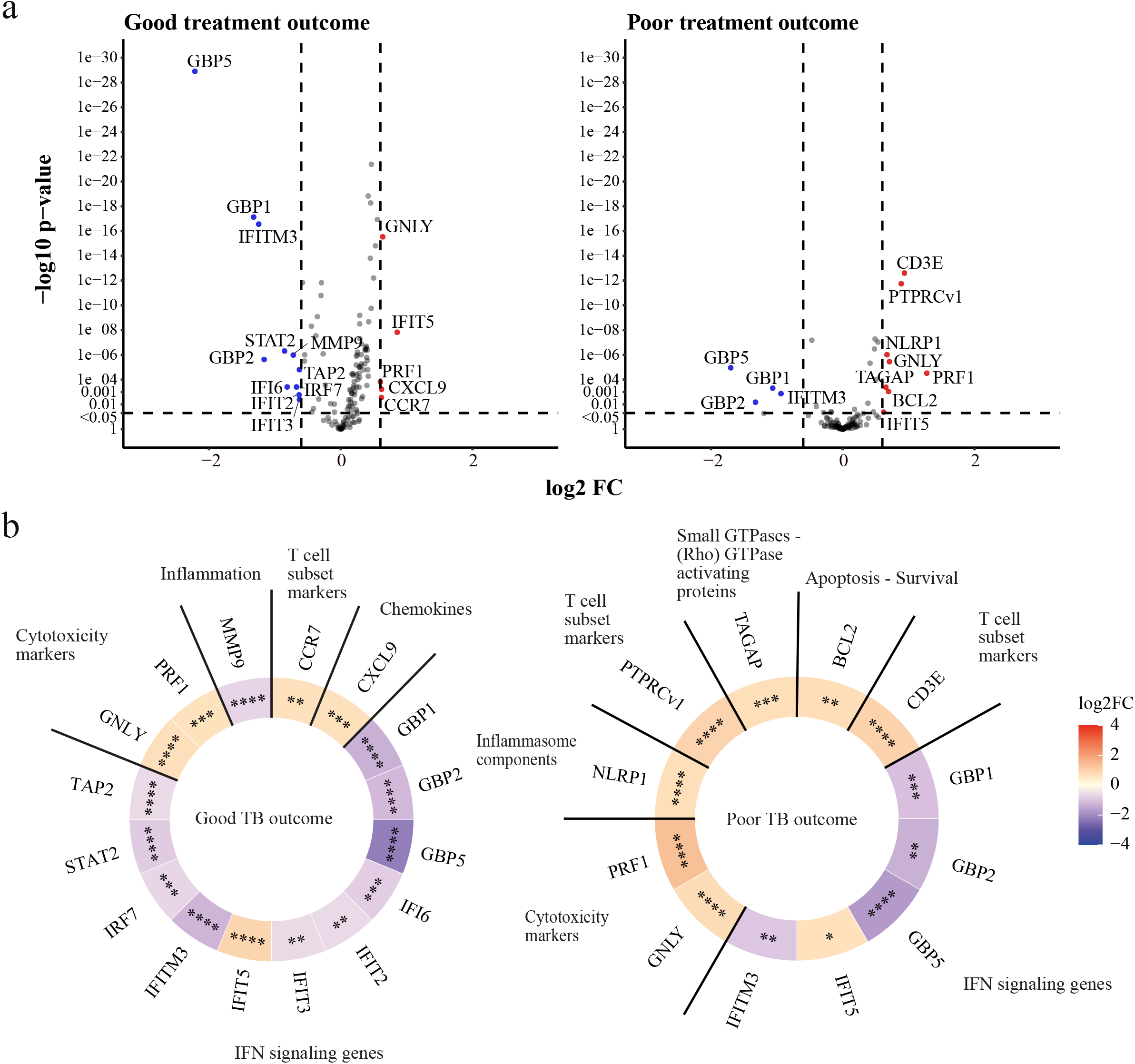
DEA of all TB patients from the pooled (South African and Indonesian) cohorts categorized based on treatment outcome compared to their gene expression levels at diagnosis. (A) Volcano plots representing DEGs regulated during anti-TB treatment of TB patients who had a good treatment outcome (left panel) or a poor treatment outcome (right panel). The y-axis scales of the plots are harmonized per treatment outcome. -log_10_- transformed p-values are plotted against log_2_ FC. Genes with p<0·05 and log_2_ FC<-0·6 or >0·6 were labelled as DEGs. (B) Heatmaps displaying log_2_ FC of the DEGs and corresponding gene modules. The saturation of color represents the magnitude of differential expression. Differences were significant by means of linear mixed models. * p<0·05, ** p<0·01, *** p<0·001, **** p<0·0001.

### Identification of a signature predicting treatment outcome

Machine learning algorithms were implemented on data obtained at each time point to develop biomarker panels to predict treatment outcomes at different stages of TB treatment. First, we aimed to identify gene signatures from RNA-Seq analysis on a subset of subjects, but we found a low performance of gene signatures generated on diagnosis, week two and month six (AUC=0·625, AUC=0·667 and AUC=0·615, respectively) to predict treatment outcome, potentially due to a low number of patients in the training and test set (Supplementary Figure S8A). The best performing model was built on month two resulting in an AUC of 0·8667 (Supplementary Figure S8A, Supplementary Table S7). We also tested an active TB disease biomarker signature, namely the three-gene Sweeney signature^20^, to determine whether it resolved significantly more in those with a good treatment outcome than in those with a poor treatment outcome. This signature has previously been shown to persist in patients with persistent lung inflammation^26^. However, in our RNA-Seq data, this signature revealed an AUC of 0·5333 (Supplementary Figure S8B) highlighting that the process behind poor treatment outcome cannot be predicted by expression of these three genes.

Next, we aimed to identify early correlates of treatment outcome by implementing machine learning algorithms on gene expression as measured by dcRT-MLPA. We focused our analysis on the identification of gene predictors at diagnosis and at week two that could possibly be used in future studies to predict the occurrence of poor or good treatment outcome before or early after anti-TB treatment initiation, first by down-sampling the good treatment outcome class. The top eight ranked genes (*GBP1, FCGR1A, STAT1, IFITM3, BCL2, CCL4, TLR9, CD274*) from the diagnosis signature were used for RF machine learning model implementation (Table 3). Excitingly, the signature had a high predictive power (AUC=0·815) to classify TB patients with a good or poor treatment outcome, already before anti-TB treatment initiation (Figure 4A). Furthermore, the gene signature showed high performance on the cohorts separately (South Africa, AUC=0·845; Indonesia, AUC=0·744). Next, we investigated whether accuracy could be improved by predicting treatment outcome after initiation of anti-TB treatment, thus measuring the early treatment response. We identified a 22-gene signature to predict treatment outcome at two weeks after initiation of anti-TB treatment (Table 3). The performance of the week two signature in predicting treatment outcome was slightly improved (AUC=0·834) compared to the diagnosis signature, especially in patients from the Indonesian cohort (AUC=0·867 versus AUC=0·744 at diagnosis). Furthermore, we identified a 14-gene month two signature, which, however, demonstrated a slightly lower accuracy in predicting treatment outcome compared to diagnosis and week two gene signatures (AUC=0·791).

**Figure 4.**
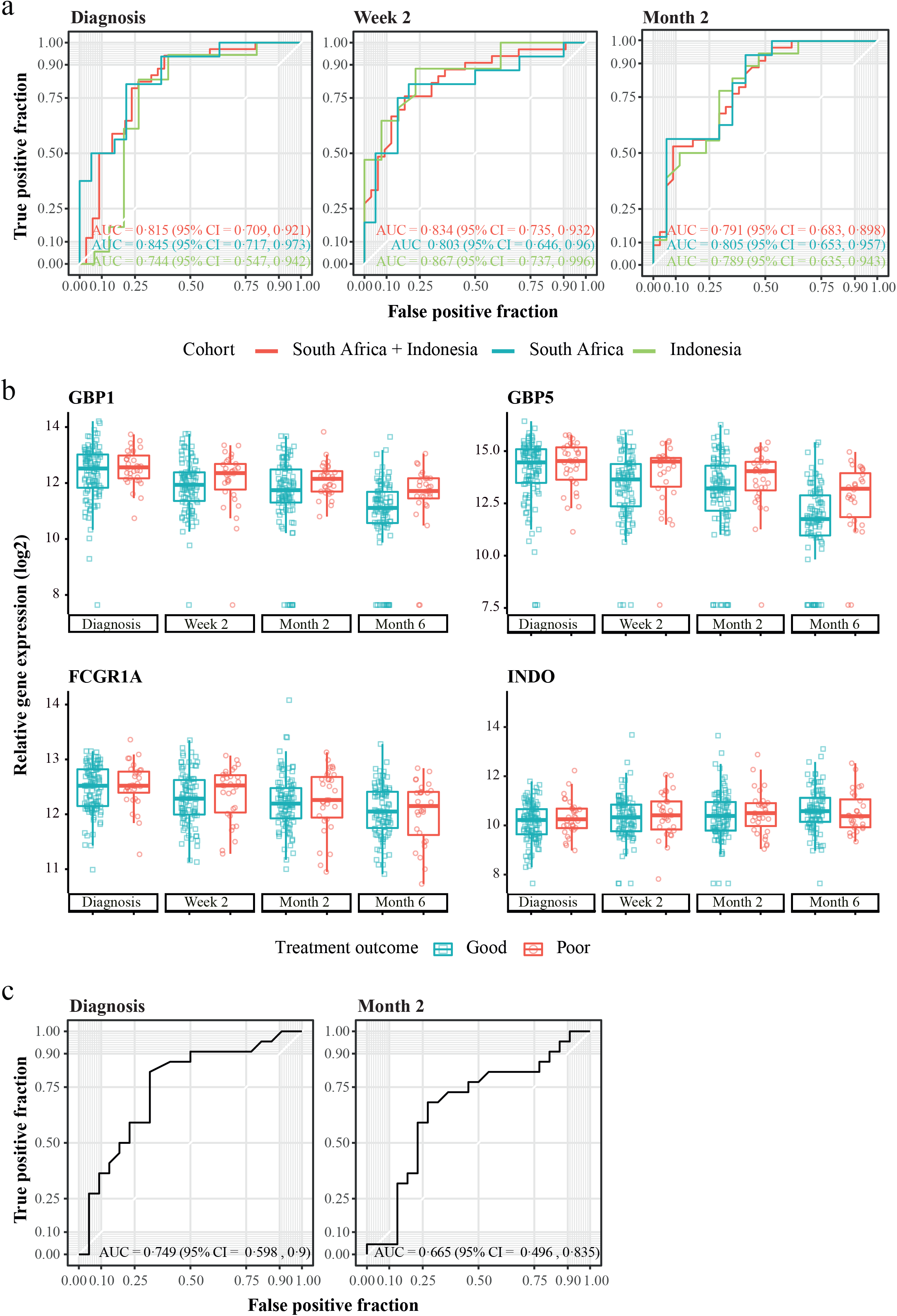
Prediction of treatment outcome in dcRT-MLPA data from peripheral blood. (**A**) ROC curves showing the predictive power of the gene signatures identified in the balanced pooled cohort (South Africa and Indonesia) to classify TB patients at diagnosis (left panel), two weeks (middle panel) or two months (right panel) after initiation of anti-TB treatment into patients who had a good treatment outcome and patients who had a poor treatment outcome, using the RFE-RF model and LOOCV. The dataset was balanced by down-sampling to encompass the same number of individuals with poor and good treatment outcome (diagnosis, n=34; week two, n=33; month two, n=34). (**B**) Gene expression kinetics of the single genes encompassing the diagnosis or week two gene signatures predicting treatment outcome in the pooled cohort. Box plots depict *GAPDH*-normalized, log_2_-transformed median gene expression values and the IQR, while the whiskers represent the data within the Q_1_-1·5xIQR and Q_3_+1·5xIQR interval. (**C**). ROC curves showing the predictive power of the gene signatures identified in the balanced pooled cohort (South Africa and Indonesia) to classify TB patients from an external validation cohort (India) at diagnosis (left panel) or two months (right panel) after initiation of anti-TB treatment into patients who had a good treatment outcome and patients who had a poor treatment outcome. The dataset was balanced by down-sampling to encompass the same number of individuals with poor and good treatment outcome (diagnosis, n=22; month two, n=22). Abbreviations: AUC, area under the curve; CI, confidence interval.

**Table 3.**
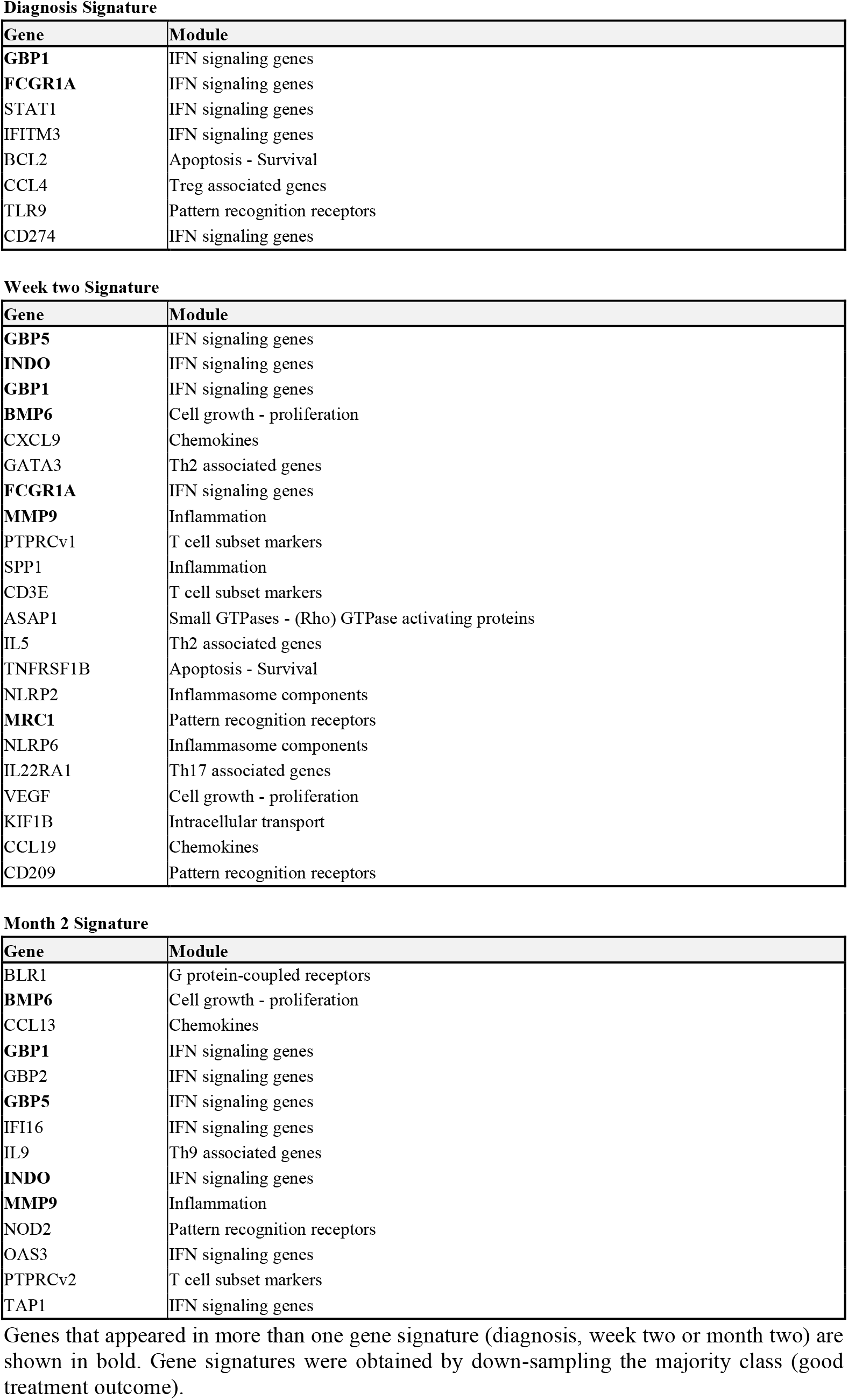
dcRT-MLPA Gene signatures Good versus Poor obtained by pooling the study groups and the cohorts

Since we detected differences in the kinetics of gene expression of patients who had a good treatment outcome versus patients with a poor treatment outcome (Figure 1), we next assessed whether a “delta” gene signature, by subtracting week two values from diagnosis, could improve the predictive performance. The delta signature encompassed seven genes (*GNLY, MRC1, GBP5, NLRP1, FLCN1, ZNF532,* and *IFIT2*) and slightly improved predictive performance (pooled cohorts, AUC=0·849; South Africa, AUC=0·839 and Indonesia, AUC=0·872) compared to the week two and diagnosis signatures (Supplementary Figure S9, Supplementary Table S8). Multiple genes were included in more than one gene signature (Supplementary Figure S10A), of which four genes (*GBP1, GBP5, FCGR1A, INDO*) are shown in Figure 4B. Next, we validated performance of the diagnosis signature and month two signature on an independent validation cohort^28^, which like our cohorts, included diabetic patients. Our diagnosis gene signature had high predictive power on the Indian validation cohort (AUC=0·749) (Figure 4C). The week two and delta signatures could not be validated on the Indian cohort, because samples were not collected two weeks after initiation of anti-TB treatment in this cohort. Three genes (*CD3E, PTPRCv1, NOD2*) that were included in our gene signatures, were also part of gene signatures described by Sivakumaran et al. (Supplementary Figure S10B). Finally, we assessed whether gene signatures with improved performance could be obtained by applying SMOTE^49^ as an alternative sampling technique. A diagnosis SMOTE gene signature was obtained that showed overlap with the diagnosis gene signatures obtained by random down-sampling (Supplementary Table S9, Supplementary Figure S10C). The SMOTE signature produced a high degree of accuracy in discriminating patients with a good treatment outcome from patients with a poor treatment outcome, but performed with lower accuracy compared to the diagnosis signature obtained by random down-sampling (pooled cohorts, AUC=0·728; South Africa, AUC=0·695; Indonesia, AUC=0·765) (Supplementary Figure S11). The diagnosis SMOTE signature exerted a similar predictive capacity on the external Indian cohort compared to the down-sampling signature (SMOTE, AUC=0·704; down-sampling, AUC=0·749).

Taken together, we identified gene signatures with high predictive power on treatment outcome, irrespective of DM as comorbidity, in patients from South Africa and Indonesia and in patients from the external Indian validation cohort.

## Discussion

In this study, we identified peripheral blood transcriptional signatures which predict anti-TB treatment success and failure in TB patients with or without concomitant hyperglycaemia or DM. Previous studies developing biomarker signatures of TB treatment success, recurrence or failure^24–26, 56^ did not include people with DM comorbidity, and we have previously found concomitant DM impairs existing TB diagnosis signature accuracy.^32^ Here we showed DM also affects existing TB treatment-response biomarker signatures in the RNA-Seq dataset, suggesting that they should be derived with cohorts including this population.

Our whole cohort dataset, from which we generated treatment outcome signatures, was derived using our dcRT-MLPA gene set, which did not contain most of the genes reported in previous signatures, except *GBP5*, which was included in our week two and month two gene signatures. Sivakumaran et al.^28^ recently reported baseline and month two gene signatures predicting treatment outcome at six months after initiation of anti-TB treatment, using the same material (whole blood), technique (dcRT-MLPA) and gene set. Notably, our treatment outcome gene signatures showed some overlap with the signatures reported by Sivakumaran et al. (*CD3E, PTPRCv1, NOD2*), suggesting that these genes are useful in predicting treatment outcome independently of ethnical background. Furthermore, our treatment outcome gene signatures showed overlap of genes of the TB risk signature predicting TB progression from healthy controls more than a year before onset of TB (*GBP1, GBP2, GBP5*, *FCGR1A, STAT1, TAP1*).^22^ Within our study, 12 genes (*BCL2, BMP6, CCL13, CD209, FCGR1A, GBP1, GBP5, INDO, MMP9, MRC1, STAT1, TLR9*) were overlapping between gene signatures, including both the gene signatures obtained by down-sampling and the gene signatures obtained by SMOTE. The occurrence of genes in multiple gene signatures within this study and between studies highlights the power of transcriptomic biomarkers in predicting treatment outcome and suggests that universal biomarkers can be applied to cohorts of different ethnicity and independently of the DM/glycaemia status of TB patients.

Patients with a poor treatment outcome responded to anti-TB treatment at the level of individual genes, as detected by downregulation of genes (*GBP1, GBP2, GBP5, IFITM3*) that have been associated with active TB and upregulation of genes (*CD3E, PTPRCv1, NLRP1, GNLY, PRF1, BCL2*) that show lower expression in patients with active TB compared to LTBI or healthy controls^15, 20, 39, 55^. However, MDP analysis showed that the response to anti-TB treatment was diminished in those with a poor treatment outcome compared to patients who had a good treatment outcome. Notably, the majority of genes that were significantly downregulated in patients who had a good treatment outcome, but not in patients who had a poor treatment outcome, are involved in IFN signaling (*IRF7, IFIT2, IFIT3, STAT2, IFI6, TAP2*). This suggests that a poor treatment outcome was reflected by persisting IFN signaling response and supports a role for type I IFN signaling in TB pathogenesis.^15, 57^

*TAGAP* was significantly increased in patients who had a poor treatment outcome in the pooled South African and Indonesian cohort as well as in both cohorts separately. *TAGAP* encodes T-cell activation Rho-GTPase-activating protein, however, the exact role of *TAGAP* in *Mtb* pathogenesis is currently unknown. Several studies have linked *TAGAP* with active TB; *TAGAP* was enriched for differential acetylation peaks upon *Mtb* infection in granulocytes^58^ and *TAGAP* was induced upon vaccination with AERAS-402 vaccine encoding a fusion protein of *Mtb* antigens.^59^ Furthermore, *TAGAP* had higher expression in TB patients compared to LTBI and healthy controls^55^ and, surprisingly, lower expression in pulmonary TB compared to household controls.^60^ Our data showing that *TAGAP* expression was significantly increased during anti-TB treatment in patients who had a poor treatment outcome could indicate that *TAGAP* is actively involved in TB pathogenesis or that *TAGAP* expression is a consequence of persisting *Mtb* infection, but this remains to be investigated.

There are several limitations of the current study. First, the sample size in this study was not based on an a priori power calculation, as this study was part of a larger study investigating differences in gene expression in patients with varying degrees of hyperglycemia. To increase statistical power, we therefore pooled patients from two cohorts (South Africa and Indonesia), which introduced heterogeneity within the studied groups. However, this can also be a strength, potentially increasing application over different ethnic backgrounds. Second, there were missing values in the cohort study. The missing values occurred as a result of random drop-outs or technical errors caused by low quantity or quality of some samples, and therefore the use of linear mixed models for the DEA most likely produced unbiased results. Third, although the prevalence of hyperglycaemia/DM is not indicated in the majority of other TB biomarker studies, which is a limitation of these studies considering the rising incidence of TB-DM comorbidity, our study contained many patients with high HbAc1 levels. Although this may have introduced a bias, the strength of this approach is that treatment outcome signatures have been developed that can be applied to patients independently of their glycaemia/DM status. Furthermore, we showed that our eight-gene diagnosis signature had a high performance (AUC=0·749) when tested on an external validation cohort in patients with a different ethnic background (India), which is striking since geographic or ethnic variations may significantly impact on the immune responses to TB.

In this study, we demonstrated the potential of gene signatures to predict treatment outcome, in a cohort including patients with concomitant DM or hyperglycaemia. Identification of a diagnosis gene signature containing only eight genes in this study, and even fewer genes in signatures reported by others^26, 27^, indicates that clinically-implementable biomarker signatures can be developed using transcriptomic-based approaches using easily accessible whole blood, and that are promising as surrogate marker for sputum culture conversion.

## Data Availability

RNA sequence data have been submitted to NCBI Gene Expression Omnibus (GEO) under accession number GSE193979. dcRT-MLPA data can be found in Supplementary Table S6.

https://www.ncbi.nlm.nih.gov/geo/query/acc.cgi?acc=GSE193979

## Contributors

Study concept and design: BA, RvC, RR, PCH, GW, SAJ, JAC, MCH, HMD, THMO, JMC; Patient recruitment, Sample collection, processing and selection: BA, KR, STM, LK, PCH, KS, RR, GW; Clinical database design, curation, maintenance: SK-B, JAC; Laboratory Experiments and data acquisition: CE, SvV, JSL, JMC; Data Analysis and interpretation: CLRvD, CE, SvV, VK, SAJ, CW, MCH, HMD, THMO, EV, JMC; Writing the manuscript: CLRvD, CE, SAJ, HMD, THMO, EV, JMC; Critical Revision of the manuscript: CLRvD, CE, KR, STM, SAJ, PCH, JAC, MCH, HMD, THMO, EV, JMC; All authors read and approved the final version of the manuscript.

## Declaration of Interests

GW had patents to methods of tuberculosis diagnosis and to tuberculosis biomarkers unrelated to the current study. The rest of the authors declare no financial or commercial conflicts of interest.

## Acknowledgements

The authors acknowledge all participants involved in this study. The authors acknowledge Bahram Sanjabi, Desiree Brandenburg-Weening, and Pieter van der Vlies for assistance with the RNA-Seq, Evelien Temminck for providing technical assistance with dcRT-MLPA experiments, Erni Durdevic for providing statistical and machine learning advice, and Prof. Dr. Harleen Grewal and Dr. Dhanasekaran Sivakumaran for providing datafiles of the cross-validation dataset.

## Supplementary Information

### Reverse-Transcriptase Multiplex Ligation-dependent Probe Amplification (dcRT-MLPA)

For each target-specific sequence, a specific RT primer was designed located immediately downstream of the left- and right-hand half-probe target sequence. 125 ng RNA was reverse transcribed to cDNA by incubation at 37°C for 15 min, using RT-primer mix and Moloney Murine Leukemia Virus (M-MLV) reverse transcriptase (Promega, Leiden, The Netherlands). Reverse transcriptase was inactivated by heating at 98⁰C for two minutes. The left- and right-hand half probes were hybridized to the cDNA at 60⁰C overnight and annealed half-probes were ligated at 54⁰C for 15 minutes using ligase-65 (MRC-Holland). Ligase-65 was subsequently inactivated by heating at 98⁰C for five minutes. Ligated probes were amplified by PCR: 33 cycles at 95⁰C for 30 seconds, 58⁰C for 30 seconds and 72⁰C for 60 seconds, followed by one cycle at 72⁰C for 20 minutes. PCR products were 1:10 diluted in Highly deionized (Hi-Di) formamide (ThermoFisher) containing 400HD Rhodamine X (ROX) fluorophore size standard (ThermoFisher). PCR products were denatured at 95⁰C for five minutes, stored immediately at 4⁰C and analyzed on an Applied Biosystems 3730 capillary sequencer in GeneScan mode (BaseClear, Leiden, The Netherlands). Trace data were analyzed using GeneMapper software 5 (Applied Biosystems, Warrington, UK). The areas of each assigned peak (arbitrary units) were exported for analysis in R (version 3.6.3). Data were corrected for batch effect and normalized to housekeeping gene glyceraldehyde 3-phosphate dehydrogenase (*GAPDH*). Signals below the threshold value for noise cutoff in GeneMapper (log2 transformed peak area 7·64) were assigned the threshold value for noise cutoff.

RT primers and half-probes were designed by Leiden University Medical Centre (LUMC, Leiden, The Netherlands) and comprised sequences for four housekeeping genes and 144 selected key immune-related genes to profile the following compartments of the human immune response (Supplementary Table S1): (1) Adaptive immune responses: T-cell responses; Th1 responses; Th2 responses; Th17/22 responses; Treg responses; T-cell cytotoxicity; Immune cell subset markers including B-cells and NK-cells. (2) Innate immune responses: Myeloid-associated markers and scavenger receptors; Pattern recognition receptors; Inflammasome components. (3) Inflammatory and IFN-signalling genes. (4) Other genes: Anti- microbial activity; Apoptosis/cell survival; E3 ubiquitin protein ligases; Small GTPases/(Rho)GTPase activating proteins; Additional chemokines; Cell growth/proliferation; Cell activation; Transcriptional regulators/activators; Intracellular transport; Mitochondrial Stress/Proteasome; Inflammation.

### Linear Mixed Models for identification of DEGs

Longitudinal DEGs were identified by means of linear mixed models using the lmer function of the lme4 package in R.^47^ To increase statistical power, datasets of all TB patients included in the South African and Indonesian cohorts were pooled independent of diabetes/glycaemia status and split based on treatment outcome. Models were fitted on *GAPDH*-normalized log_2_- transformed targeted gene expression data. Outcome-time interactions were included as fixed effects and the patients ID-time interactions were included as random effects.

### Identification of gene signatures for treatment outcome

For modelling analyses, RNA-Seq data were randomly split into training and test sets (60/40) using the R package *caret*.^51^ Feature selection was performed for each timepoint using RFE^48^ with repeated cross validation as the re-sampling method (n=10). A weighted model was fitted using glmnet method, using weights 1/frequency * 0·5 and repeated cross validation for re- sampling (n=10). Each model was used to make predictions on the corresponding test set.

To identify signatures associated with treatment outcome in dcRT-MLPA data, TB patients of South Africa and Indonesia were pooled independent of diabetes/glycaemia status. To balance the dataset, we applied two random sampling approaches using R: (1) a down- sampling approach, reducing the number of patients in the majority class (i.e. good treatment outcome) and (2) an up-sampling approach by generating synthetic data from existing data using the SMOTE^49^ function from the *DMwR* package in R, resulting in equal numbers of patients in both classes. Then, RFE^48^, available in the *caret* R package^51^, was applied with K-fold validation (K=10) to the entire data set search for the optimal combination and number of top-ranking genes able to separate TB patients with a good and poor treatment outcome. RFE is a powerful approach for variable selection in high-dimensional data by selecting features that fit a model and removing the weakest feature (or features) until the specified number of features is reached. Once the best predictors of treatment outcome were identified, the expression values of these genes were extracted from the dataset. We subsequently applied RF^50^ as machine learning algorithm on the dataset and evaluated the performance by LOOCV, both available on the *caret* R package.^51^

---

**Supplementary Figure S1.**
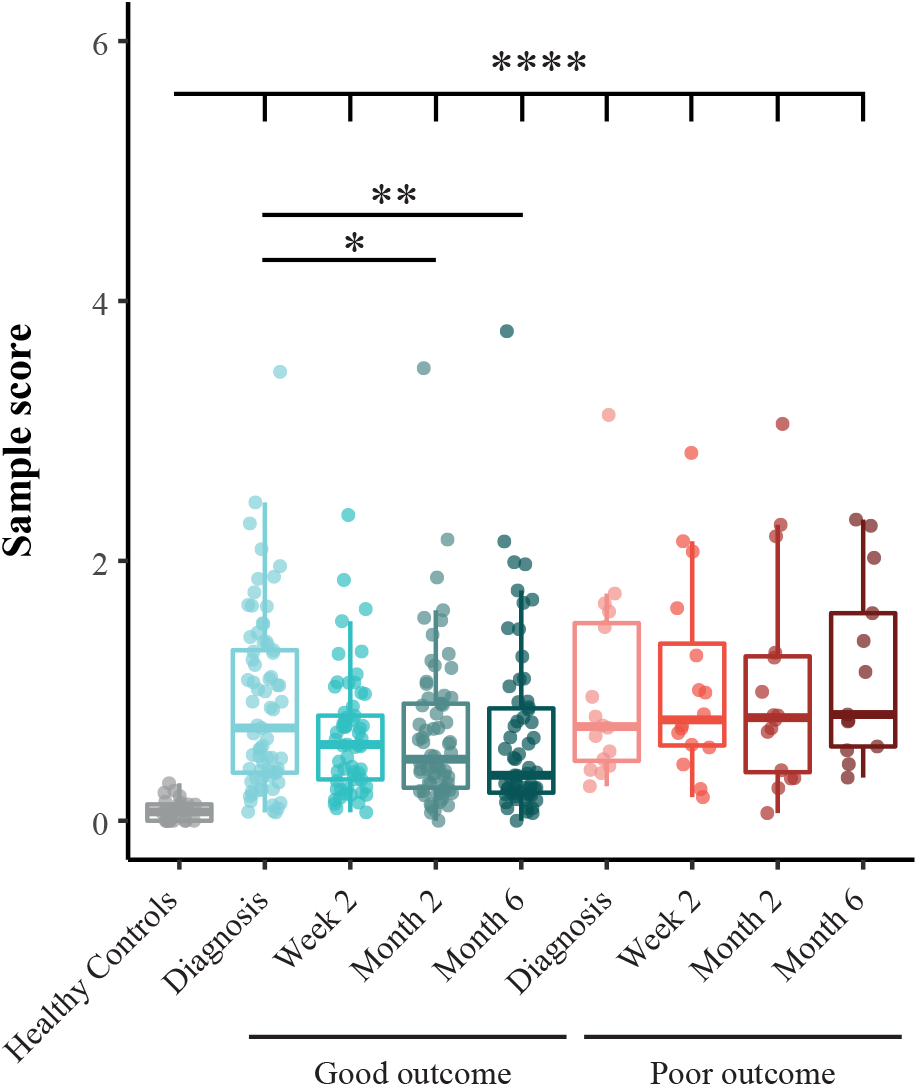
MDP plot representing the change in gene expression perturbation in TB patients from South Africa categorized based on treatment outcome. Blood transcriptomes from TB patients who had a good or poor treatment outcome were determined by dcRT-MLPA. The extent of overall difference in gene expression, relative to the median of expression in healthy controls, was calculated for individual patients at the timepoints shown. The bars and whiskers show the median and data within the Q1-1·5 x inter quartile range (IQR) and Q3+1·5 x IQR interval. Differences were significant by Mann-Whitney U-test with Benjamini-Hochberg correction for multiple testing. * p<0·05, ** p<0·01, *** p<0·001, **** p<0·0001.

**Supplementary Figure S2.**
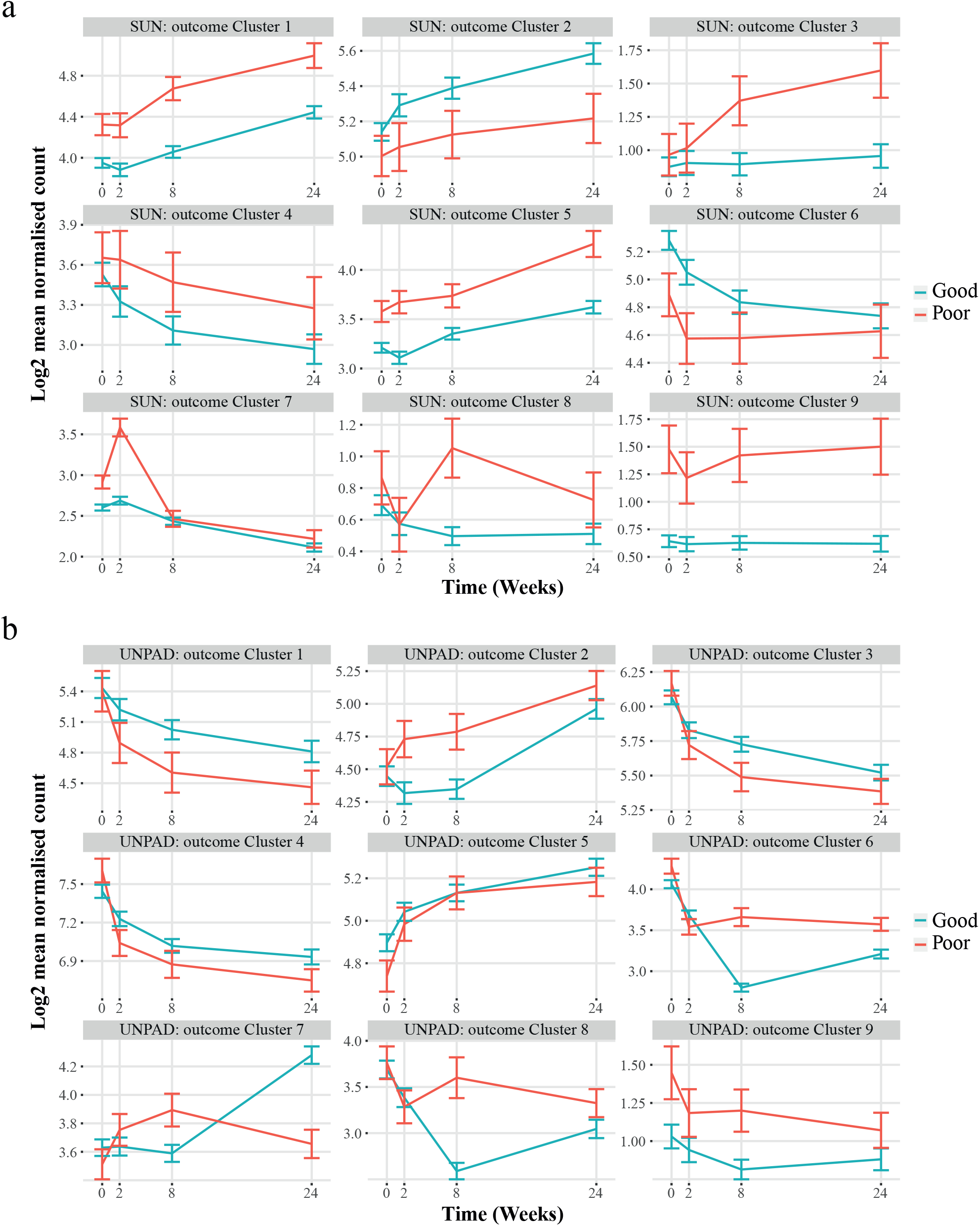
Differential change in gene expression in TB patients through treatment in those who had a good or poor treatment outcome. MaSigPro analysis was conducted on the blood RNA-Seq data from TB patients from South Africa (a) or Indonesia (b), to identify genes which were significantly differentially expressed between those patients with a good or poor outcome. Plots show hierarchical clusters of genes, and bars show mean ± 1 SEM. Data were filtered to remove lowly abundant transcripts prior to analysis.

**Supplementary Figure S3.**
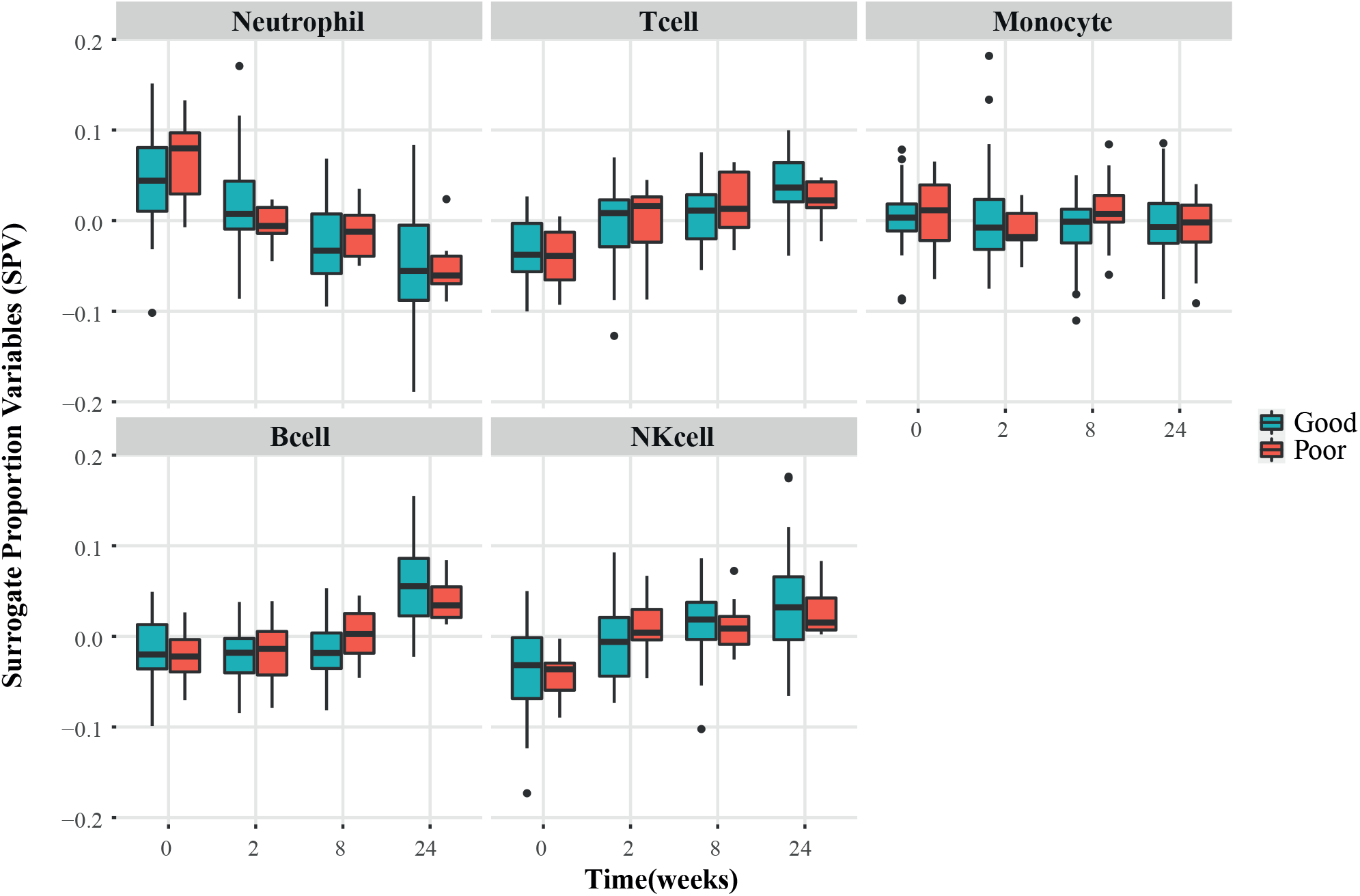
Cell population estimates in good and poor treatment outcomes in South Africa and Indonesia. Estimates of relative differences in cell proportions were calculated from RNA-seq data using R package CellCODE. IRIS and DMAP data sets used as reference. Bars and whiskers show median and 1·5 x IQR.

**Supplementary Figure S4.**
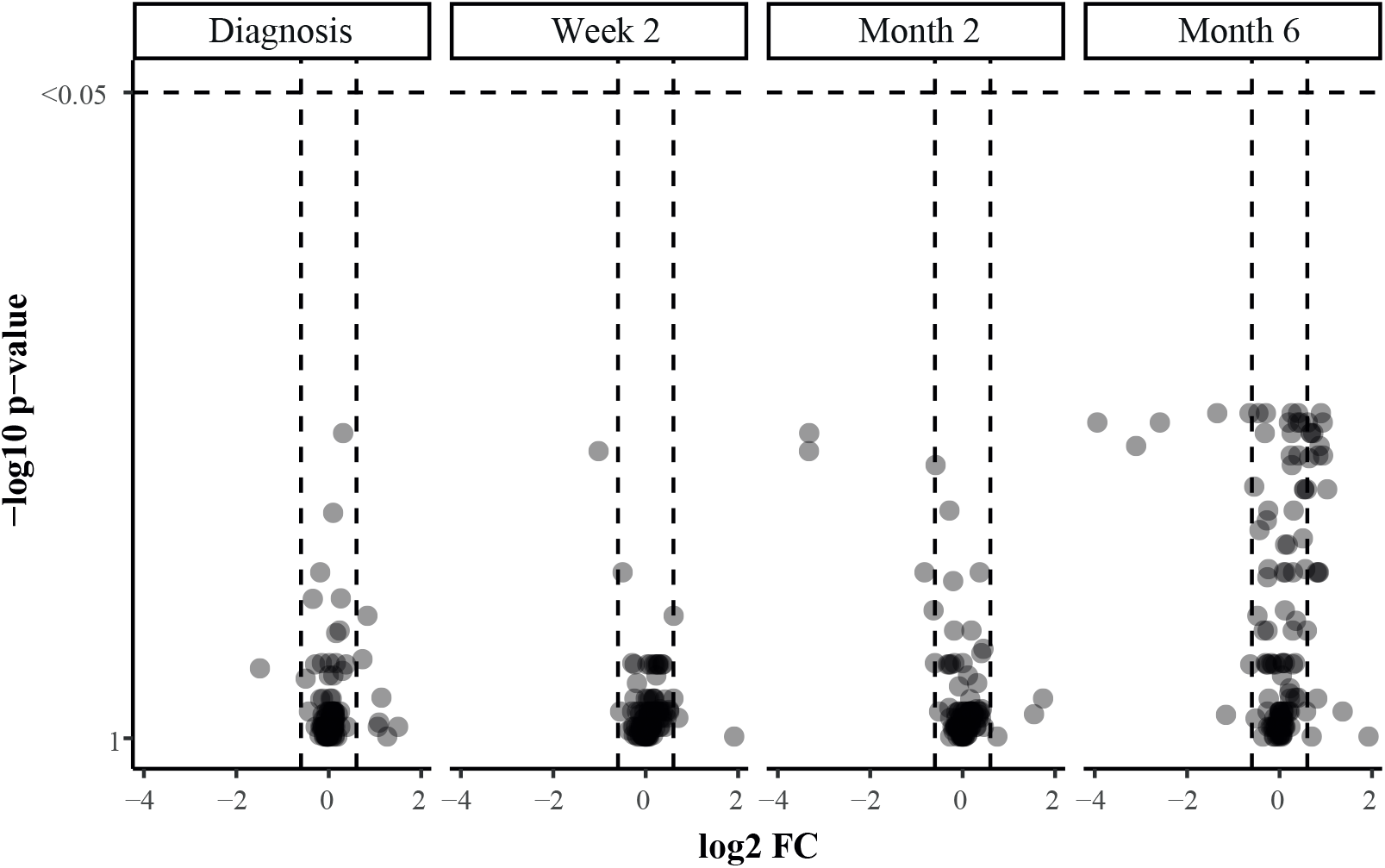
Differential expression analysis in patients from South Africa and Indonesia who had a poor treatment outcome versus patients who had a good treatment outcome at the indicated timepoints. Non-parametric Mann-Whitney U-test with Benjamini-Hochberg correction for multiple testing was applied to test for statistical differences between the groups.

**Supplementary Figure S5.**
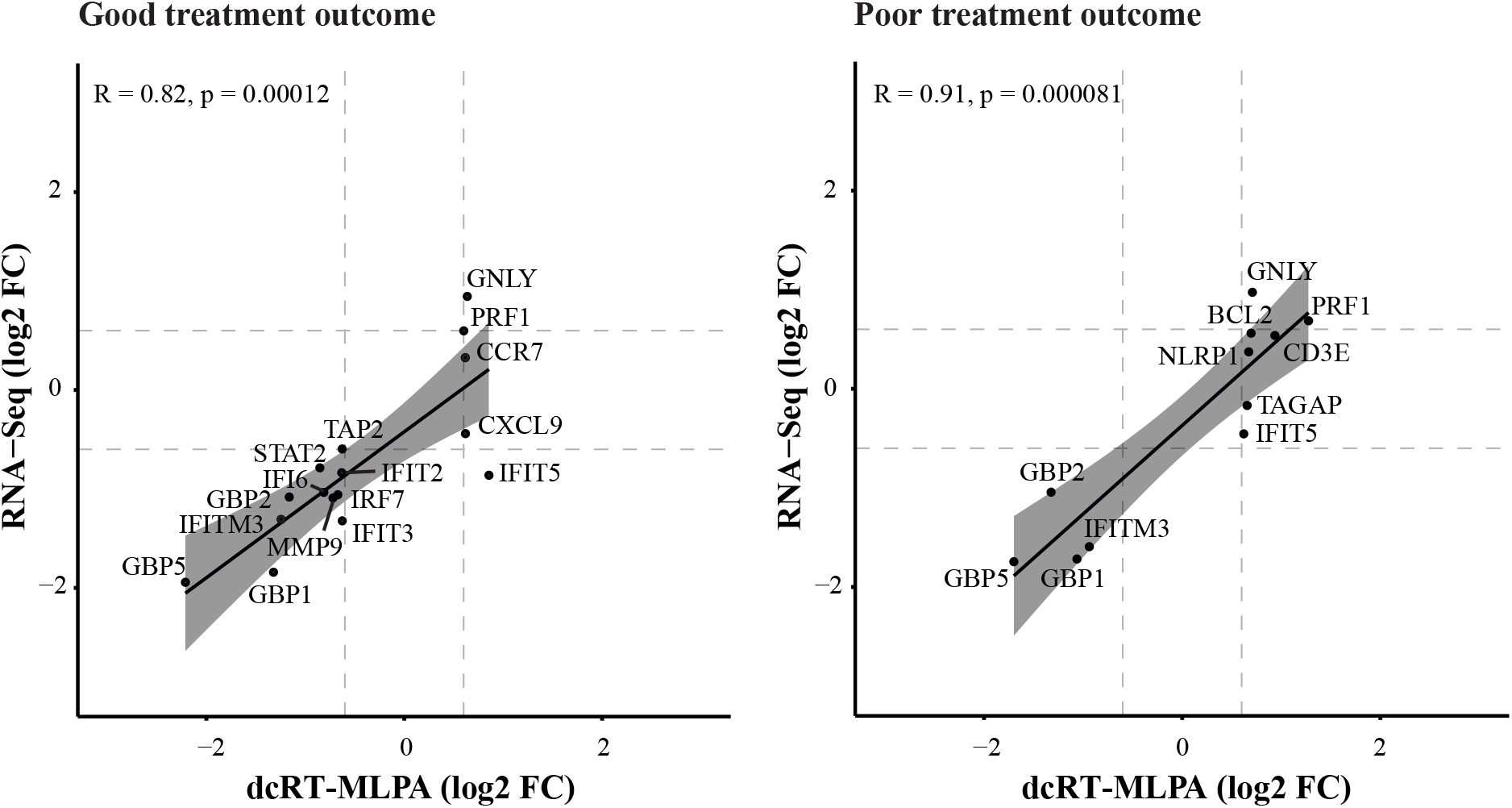
Scatter plots representing Pearson correlations between the longitudinal DEGs identified by dcRT-MLPA versus the same genes identified by RNA-Seq. Values are plotted as log2 FC (month six-diagnosis). Black line corresponds to line of best fit and shaded bands indicate confidence intervals.

**Supplementary Figure S6.**
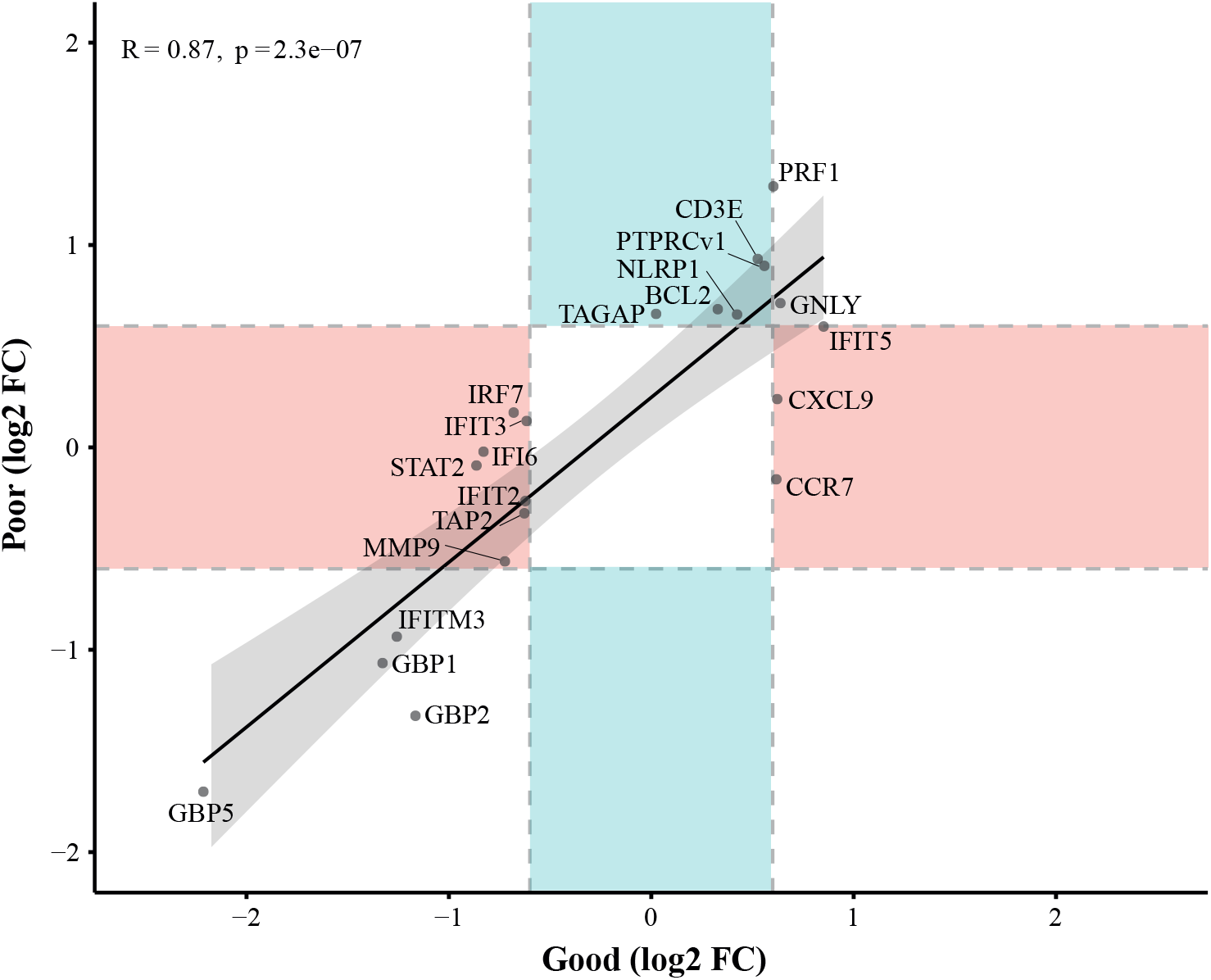
Scatter plot representing Pearson correlations between the longitudinal DEGs in TB patients who had a poor treatment outcome versus the longitudinal DEGs in TB patients who had a good treatment outcome. Values are plotted as log2 FC (month 6-diagnosis). Black line corresponds to line of best fit and shaded bands indicate confidence intervals. Red shaded areas indicate genes that were identified as DEGs only in patients who had a good treatment outcome and blue shaded areas indicate genes that were identified as DEGs only in patients who had a poor treatment outcome.

**Supplementary Figure S7.**
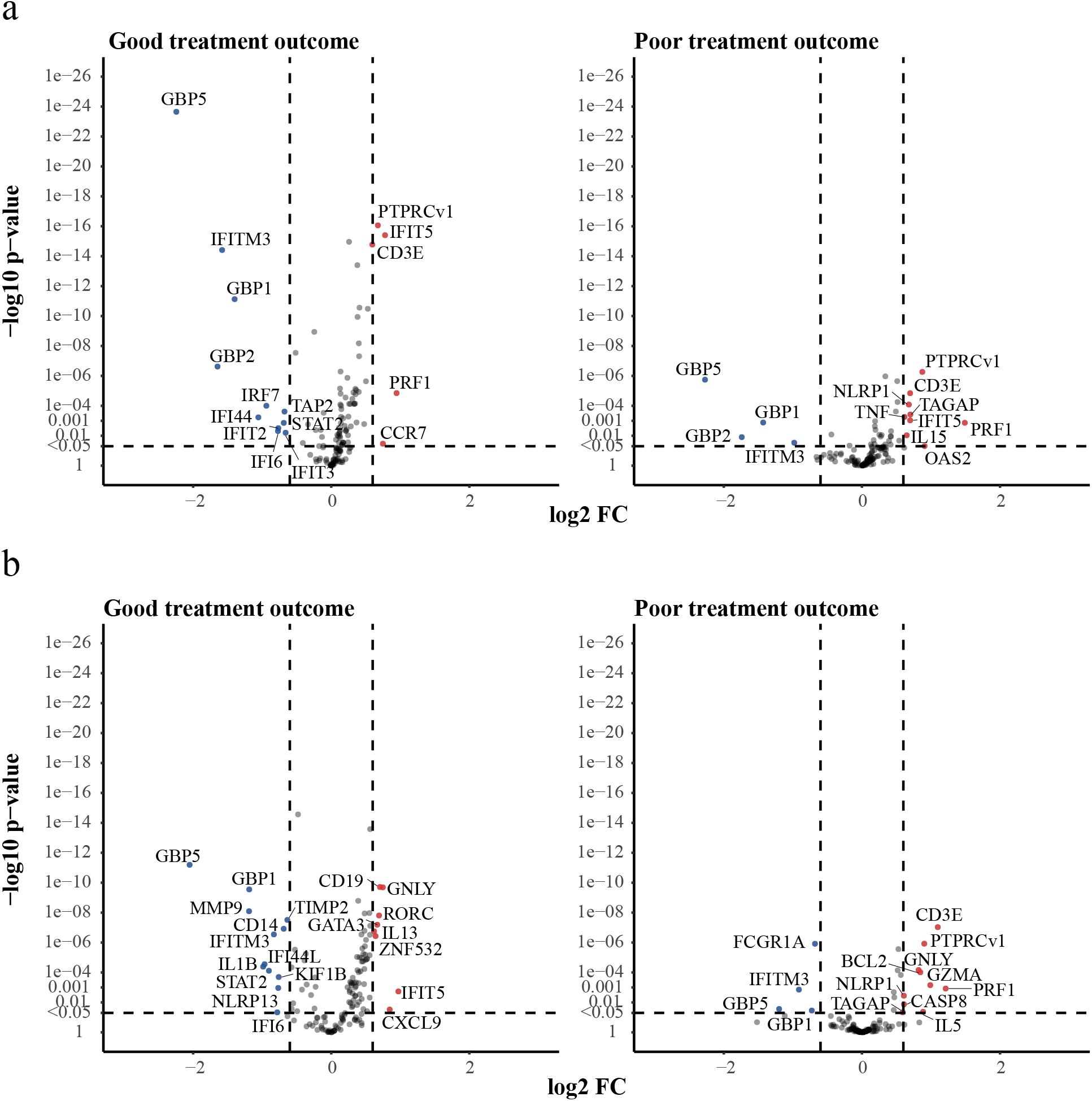
Differential expression analysis of all TB patients from the South African and Indonesian cohorts categorized based on treatment outcome compared to their gene expression levels at diagnosis. (a) Volcano plots representing DEGs regulated during anti-TB treatment of TB patients from the South African cohort who had a good treatment outcome (left panel) or a poor treatment outcome (right panel). (b) Volcano plots representing DEGs regulated during anti-TB treatment of TB patients from the Indonesian cohort who had a good treatment outcome (left panel) or a poor treatment outcome (right panel). (a,b) The y-axis scales of the plots are harmonized per treatment outcome. -log10-trans- formed p-values are plotted against log2 FC. Genes with p <0·05 and log2 FC <-0·6 or >0·6 were labelled as DEGs.

**Supplementary Figure S8.**
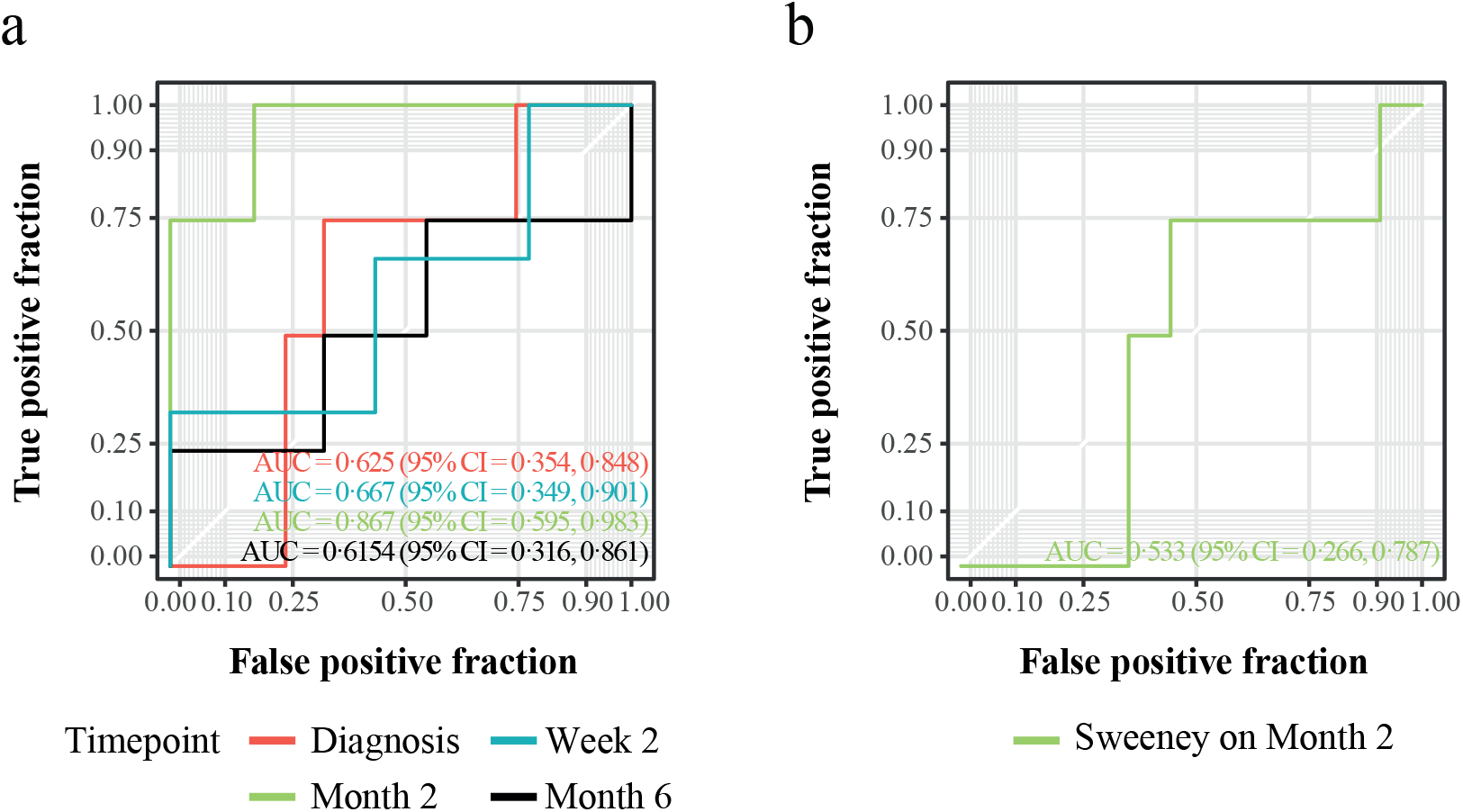
Prediction of treatment outcome in RNA-Seq data from peripheral blood. ROC curves showing the predictive power of (a) the gene signatures identified in the pooled cohort (South Africa and Indonesia) or (b) the Sweeney gene signature to classify TB patients at the indicated timepoints after initiation of anti-TB treatment into patients who had a good treatment outcome and patients who had a poor treatment outcome. Data were split into training and test sets (60/40). For each time point a gene signature was generated by RFE and a weighted model fitted using glmnet method. Weights of 1/frequency * 0.5 were used. Abbreviations: AUC, area under the curve; CI, confidence interval.

**Supplementary Figure S9.**
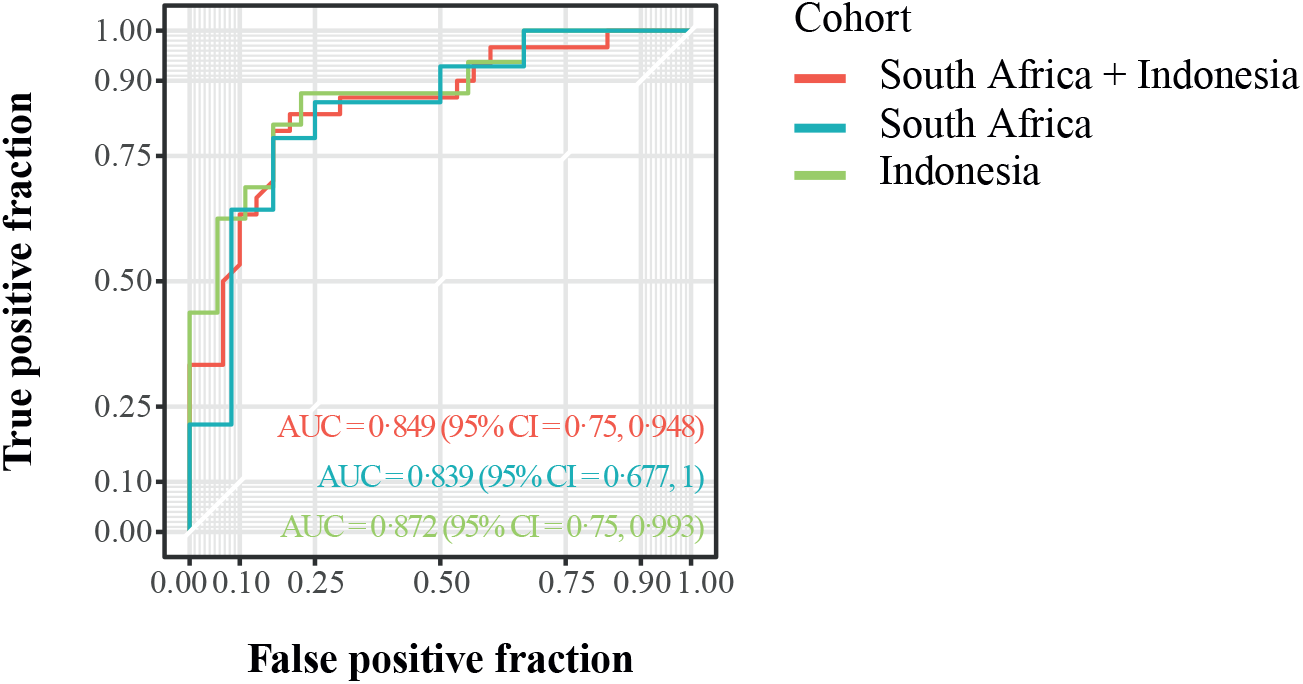
Identification of a delta (week two minus diagnosis) signature predicting the outcome of anti-TB treatment. ROC curve showing the predictive power of the gene signature identified in the balanced pooled cohort to classify TB patients into patients who had a good treatment outcome and patients who had a poor treatment outcome, using the RFE - RF model and LOOCV. The dataset was balanced by down-sampling to encompass the same number of individuals with poor and good treatment outcome. Abbreviations: AUC, area under the curve; CI, confidence interval.

**Supplementary Figure S10.**
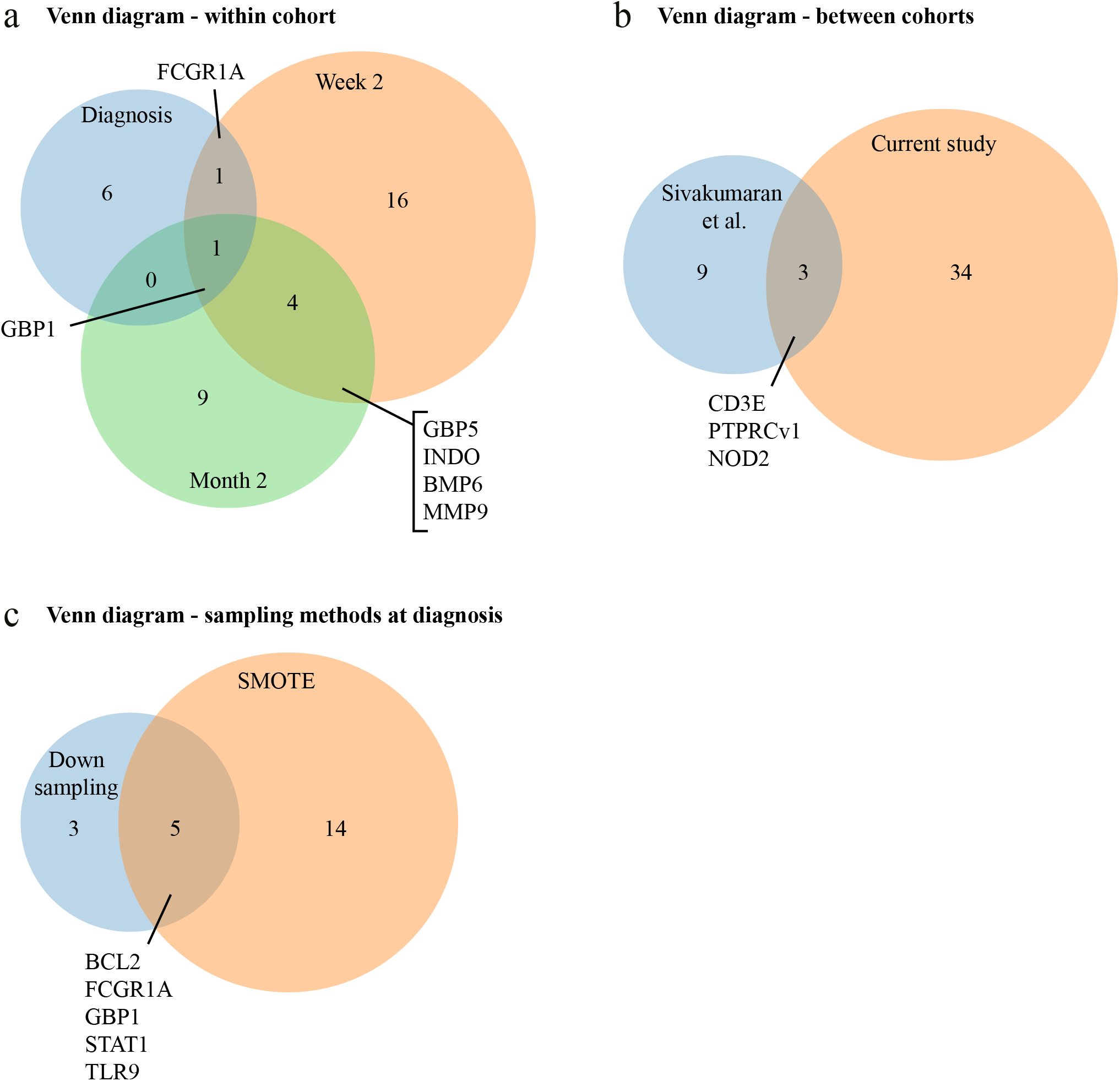
Venn diagrams showing the number of genes encompassing treatment outcome signatures as identified by RFE-RF models. (a) Venn diagram displaying the number of unique and overlapping genes comparing diagnosis (blue), week two (orange) and month two (green) gene signatures obtained by random-downsampling. (b) Venn diagram displaying the number of unique and overlapping genes comparing the gene signatures in the current study (diagnosis, week two and month two; orange) obtained by random-downsampling with the gene signatures published by Sivakumaran et al. (blue). (c) Venn diagram displaying the number of unique and overlapping genes comparing the diagnosis gene signature obtained by random-downsampling (blue) with the diagnosis gene signature obtained by applying SMOTE sampling (orange) to balance the classes. Overlapping genes are annotated.

**Supplementary Figure S11.**
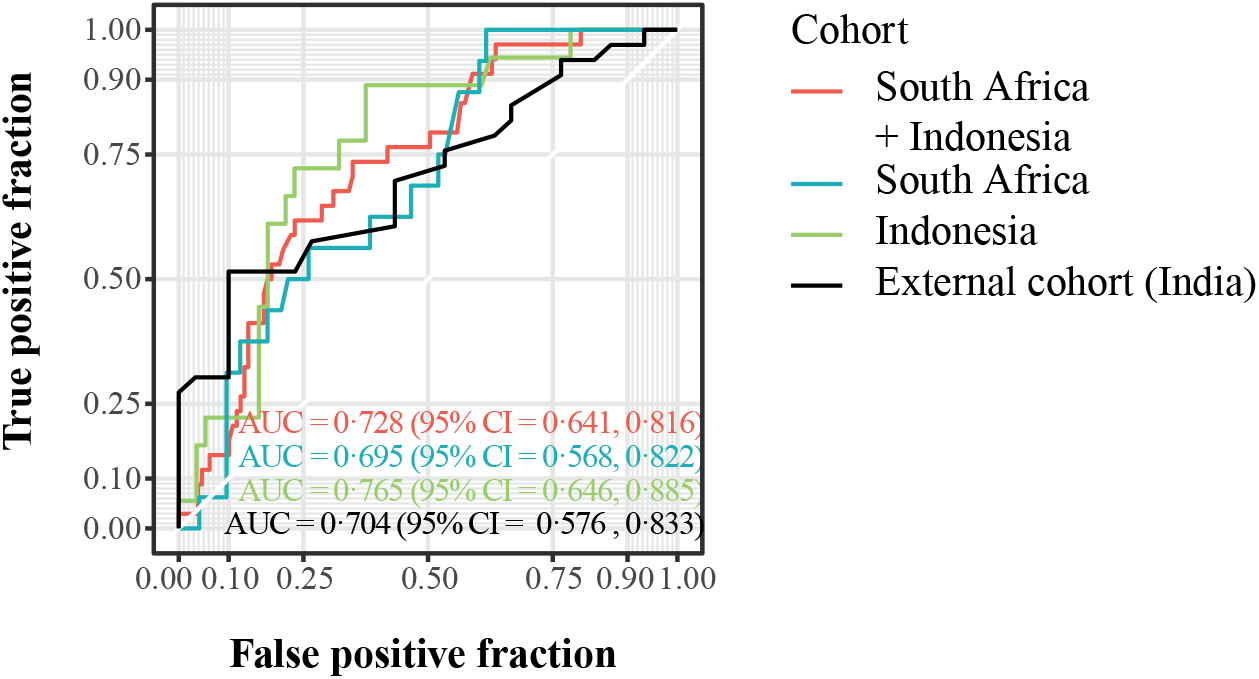
Identification of a SMOTE diagnosis signature predicting the outcome of anti-TB treatment. ROC curves showing the predictive power of the gene signatures identified in the balanced pooled cohort (South Africa and Indonesia) and validated in the South African and Indonesian cohort or in an external Indian cohort. TB patients are classified into patients who had a good treatment outcome and patients who had a poor treatment outcome at diagnosis, using the RFE - RF model and LOOCV. The dataset was balanced by SMOTE to encompass the same number of individuals with poor and good treatment outcome. Abbreviations: AUC, area under the curve; CI, confidence interval.

**Supplementary Table S1.**
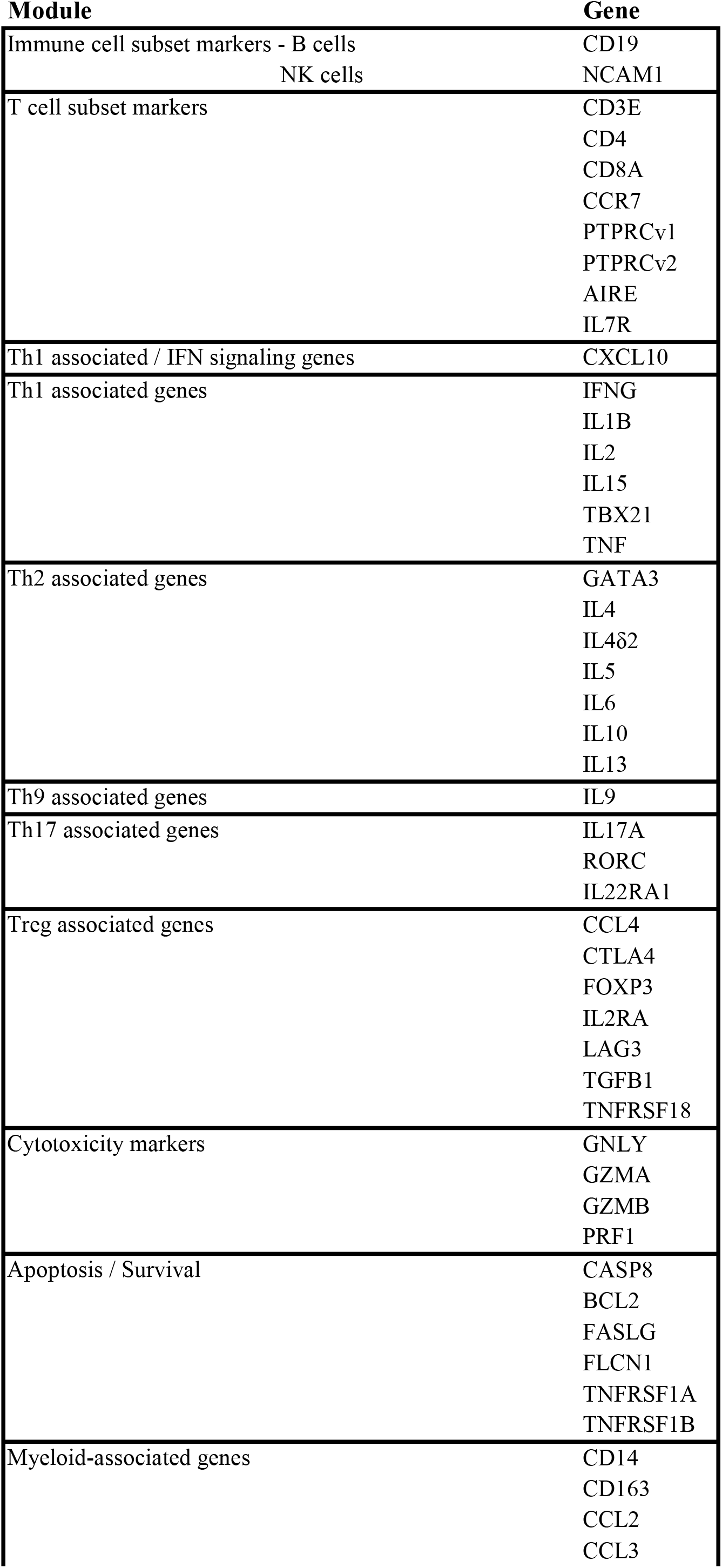
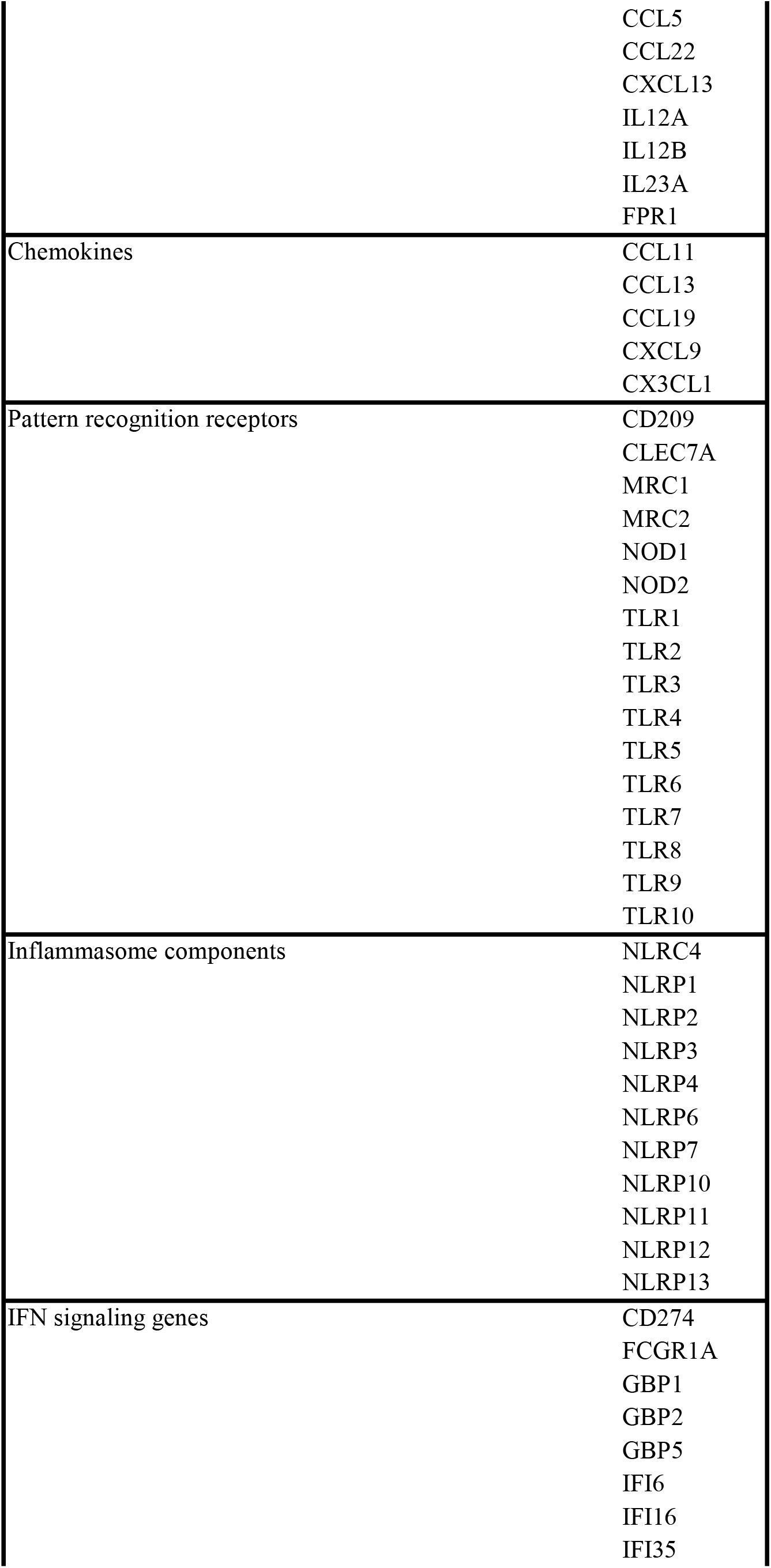
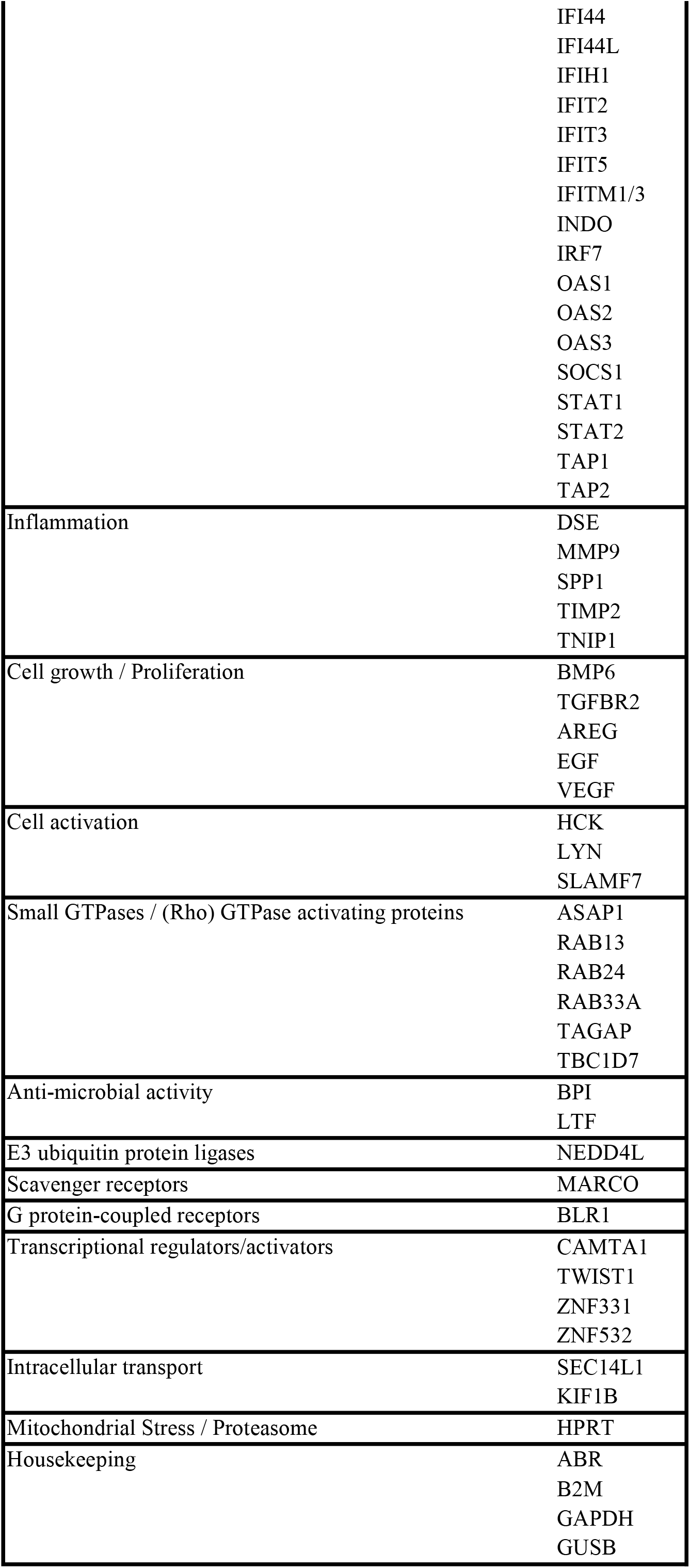
List of target genes for dcRT-MLPA.

**Supplementary Table S7.**
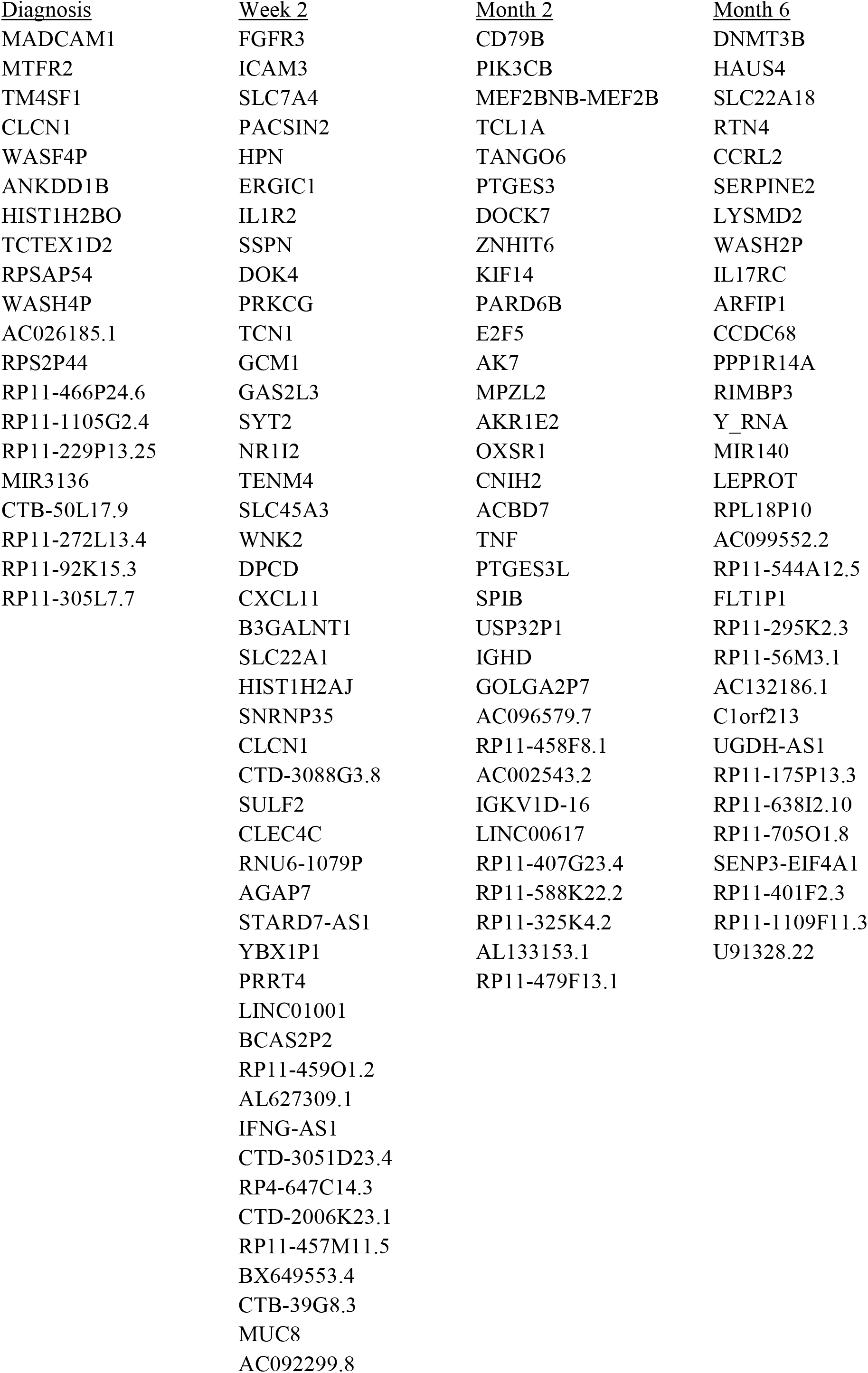
Gene signatures for each timepoint - RNA-Seq

**Supplementary Table S8.** Delta signature Good vs Poor obtained by pooling the study groups and the cohorts.

**Delta (Week two - Diagnosis) Signature**
| Gene | Module |
| --- | --- |
| GNLY | Cytotoxicity markers |
| **MRC1** | Pattern recognition receptors |
| **GBP5** | IFN signaling genes |
| NLRP1 | Inflammasome components |
| FLCN1 | Apoptosis - Survival |
| ZNF532 | Transcriptional regulators - activators |
| IFIT2 | IFN signaling genes |

Genes that appeared also in the diagnosis or week two gene signatures are shown in bold. Gene signatures were obtained by downsampling the majority class (Good treatment outcome).

**Supplementary Table S9.** SMOTE signature Good vs Poor obtained by pooling the study groups and the cohorts.

**Diagnosis Signature**
| Gene | Module |
| --- | --- |
| AREG | Cell growth - proliferation |
| **BCL2** | Apoptosis - Survival |
| CASP8 | Apoptosis - Survival |
| CCL13 | Chemokines |
| CD209 | Pattern recognition receptors |
| CLEC7A | Pattern recognition receptors |
| CTLA4 | Treg associated genes |
| CX3CL1 | Chemokines |
| **FCGR1A** | IFN signaling genes |
| **GBP1** | IFN signaling genes |
| IL12B | Myeloid associated genes |
| LTF | Anti-microbial activity |
| NLRP3 | Inflammasome components |
| **STAT1** | IFN signaling genes |
| TIMP2 | Inflammation |
| TLR8 | Pattern recognition receptors |
| **TLR9** | Pattern recognition receptors |
| TNFRSF1A | Apoptosis - Survival |
| ZNF331 | Transcriptional regulators - activators |

Gene signature obtained by applying SMOTE sampling technique. Genes that are also identified in the Diagnosis signature obtained by downsampling are shown in bold.

